# The Immunity Paradox of Bed Nets: Why Reducing Exposure Can Still Strengthen Malaria Control

**DOI:** 10.64898/2026.07.04.26355807

**Authors:** Arsène Jaurès Ouemba Tassé, Bime M. Ghakanyuy, Hemaho B. Taboe, Gideon A. Ngwa, Calistus N. Ngonghala

## Abstract

Insecticide-treated bed nets (ITNs) are central to malaria control. They serve as physical barriers and chemical agents that deter and kill mosquitoes, thereby reducing transmission; however, this form of protection reshapes the immunology of malaria by reducing exposure to *Plasmodium* parasites and weakening the development of naturally acquired immunity. Against this background, the present study develops a modeling framework to investigate how this tension between protection and immunity alters malaria dynamics once vaccination is introduced as a complementary control strategy and the optimum combination of ITN and vaccine coverage required for malaria control. Unlike standard models that use a fixed proportional reduction in transmission, this study models ITN coverage and efficacy as coupled, time-dependent processes and immunity driven by exposure to infection and vaccination. Rigorous analysis of the model identifies existence conditions for equilibria and shows that malaria can be contained through the synergistic interaction of vaccination, vector control, and immunity-mediated host dynamics. Parameter values of the model are estimated by fitting the model to confirmed malaria case data and the estimated baseline reproduction number using these parameter values is 1.41 (95% confidence interval: 1.34-1.48), confirming sustained transmission. Simulations of the model show that, although ITNs reduce immunity acquisition, their net effect is to reduce infections and improve recovery and survival. Hence, population-level benefits of ITNs outweigh their immunity-reducing effects (particularly when combined with vaccination), leading to a reduced malaria burden. Comprehensive sensitivity analysis indicates that malaria burden is driven mostly by mosquito biting intensity, population capacity, and transmission probabilities; while mosquito mortality, effective treatment, ITN performance, and vaccine efficacy cause the most significant reductions. Additionally, malaria is uncontrollable with universal ITN use and vaccination at baseline efficacy. While individual interventions can achieve control under low transmission, neither 100% ITN coverage nor 100% vaccine coverage can achieve control under moderate-to-high transmission. However, a strong synergy between ITNs and vaccination allows combinations of high efficacy to achieve containment at realistic coverage levels, suggesting that integrated malaria control involving effective vector control, vaccination, and prompt treatment is needed.

## 1. Introduction

Malaria remains a persistent challenge to public health globally, especially in Sub-Saharan Africa, South America, Southeast Asia, and Western Pacific regions, with nearly half a million deaths annually [1]. This situation is worsened in Africa owing to excessive poverty, minimal access to health care, poor infrastructure, and non-optimally implemented preventive methods. Malaria is caused by the *Plasmodium* parasite and transmitted to humans through the bite of an infected female *Anopheles* mosquito that requires blood as a nutrient source to complete the development of her eggs. Mosquitoes can serve as carriers for the disease for their lifespans [2]. The transmission cycle involves humans becoming infected when bitten by female mosquitoes, which in turn infect mosquitoes when the mosquitoes feed on infectious humans, thus, making them potential carriers of the disease to infect other humans.

Vaccination of humans against malaria is a new control strategy in addition to other interventions implemented. Nonetheless, vaccination of children is a significant intervention as malaria exhibits a higher burden in young children within endemic regions. The two recommended vaccines (RTS,S/AS01 and R21/Matrix-M malaria) at the time of writing provide partial protective benefits against developing clinical malaria and severe malaria disease upon infection by *Plasmodium falciparum* because they prevent its establishment inside hepatocytes [3]. The RTS,S/AS01 and R21/Matrix-M vaccines are delivered through a four-dose schedule, starting from around 5 months of age, and a three-dose primary series with a boost uptake, respectively, forming part of integrated routine childhood immunization programs in high transmission settings [4]. The vaccine-induced protection was moderate and progressively waned over time. However, the boost led to better protection longevity and impact [5, 6]. Early large-scale deployment and pilot programs demonstrated a significant effect in lowering malaria incidence, severe disease, hospitalizations, and mortality [7]. The impact of broad coverage of vaccines is being assessed in ongoing trials in older children and adults, with optimized delivery strategies. This is helpful in malaria control when integrated with core interventions like insecticide-treated nets, indoor residual spraying, seasonal malaria chemoprevention, and prompt case management. In particular, this will lead to even further progress toward eradication.

The human-mosquito contact interactions focus on reducing contacts between the human population and the mosquito. Some strategies promoting the minimization of malaria transmission are door and window screens, indoor residual spraying, peridomestic space spraying, insect repellents, and insecticide-treated bed nets (ITNs) [8, 9]. The term ITN applies generally and includes conventional ITNs, any long-lasting insecticidal nets, piperonyl butoxide nets, and combinations like pyrethroid–chlorfenapyr (the next generation). ITNs have become highly adopted and effective interventions in the fight against malaria transmission over the last decades, particularly in low-resource areas of endemic regions, to protect vulnerable populations from malaria [10, 11]. They provide physical blocking from mosquito bites and act as a chemical barrier by either repelling or killing them using insecticide-treated fibers. Despite these proven benefits in lowering malaria cases, their universal use has increased worries about the long-term effect on naturally acquired immunity.

Immunity is a body’s capability to resist infections either by eliminating pathogens or neutralizing their pathogenic effects [12]. In this context, immunity can build up after exposure to the malaria causative agent (the *Plasmodium* parasite). More so, in endemic areas where continued exposure builds acquired immunity to malaria, which does not prevent infection but lower parasite load, thereby lessening the severity of the disease and the likelihood of death from it [13]. In particular, simultaneous reduction in malaria incidence by ITNs also arouses reduced natural immunity for less frequent and milder cases later in life. In this regard, most individuals, including children, in extreme endemic regions continue to remain immunologically naive or lose acquired protection and thereby become susceptible to severe malaria. Therein lies the critical conundrum: while the use of ITNs is effective in lowering malaria transmission and associated disease burden, it requires the development of acquired immunity over time at the population level. As such, the long-term effects on immunity and, hence, the severity of malaria make this situation non-intuitive. Hence, it is important to assess not only the influence of ITNs on the dynamics of malaria transmission but also immunity build up and disease severity. Understanding this balance is crucial for guiding integrated malaria control strategies that combine ITNs with complementary interventions, such as vaccination and other vector control measures, to maximize public health benefits while minimizing unintended risks.

A variety of mathematical models have been developed to understand the dynamics of malaria transmission [14– 26]. Several of these studies focused on the role of ITNs in reducing malaria cases [17, 18, 27–29]. In [17], the authors established that bed nets have a substantial positive effect in reducing the basic reproduction number, thereby alleviating the malaria burden. They concluded that if 75% of the population consistently use bed nets, malaria could be eliminated. Building on this, [30] refined the analysis by distinguishing between individuals who own bed nets and those who use them correctly, and by dividing the mosquito population into resting and questing groups. Through optimal control theory, they showed that although bed net use may reduce immunity levels, this drawback is outweighed by the reduction in disease transmission. However, they highlighted that improper use of ITNs remains a critical threat. The studies in [18, 27–29] further investigated ITN effectiveness by incorporating the effects of physical and chemical degradation of nets over time, alongside human behavioural factors, insecticide resistance, and temperature variation. The results show that the basic reproduction number and infection burden are most sensitive to ITN coverage and mosquito biting rates. Also, incorporating asymptomatic infection, insecticide resistance, ITN decay, and temperature-driven seasonality reveals underestimation of malaria risk, and complex transmission dynamics, underscoring the need for high-quality long-lasting ITNs, timely replacement, and temperature-aware control strategies. Another study described in [31] incorporates population immunity boosted by repeated exposure and its role in generating asymptomatic carriers who sustain transmission. Results of the study show that heterogeneous mosquito biting shapes immunity acquisition and disease dynamics and highlight the fact that while vector control reduces transmission, it may also lessen acquired immunity, implying that immunity-aware strategies are essential for effective and sustained malaria control.

To our knowledge, none of the reported research items has interrogated the apparently paradoxical protection that comes with ITN use explicitly. Therefore, we pose the following questions: Is the scenario whereby sustained use of ITNs to protect the user against repeated malaria infections, which may be seen as a pathway to strip the user off the much needed opportunity to build up naturally acquired immunity to malaria through repeated exposure to the disease, eventually detrimental to the ITN user? If naturally acquired immunity to malaria is sustained by continuing exposure, and blocking sustained transmission via use of ITNs means acquired immunity wanes, what, therefore, is the impact and interplay between sustained ITN-use, vaccination, and immune status? This paper answers the questions by formulating and studying a model that accounts for the transmission dynamics of malaria, the long-term impact of ITN usage, vaccination, and immunity build-up. This is achieved by accounting for the following underlying mechanisms: 1) ITN coverage and efficacy as dynamic state variables; 2) the effect of reduced mosquito feeding and population, as well as naturally acquired immunity build-up due to ITN-use; and 3) the build-up and decay dynamics of human immunity through repeated exposure to malaria infection and vaccination. This differs from traditional modeling in which intervention through ITN-use is treated as a constant modifier on transmission. This dynamic representation captures efficacy decay and replacement-driven renewal, enabling realistic transient and long-term intervention effects that static reduction models cannot capture. The framework is used to assess whether the use of ITNs is beneficial or detrimental to malaria control since it reduces acquired immunity and to determine the optimum ITN and vaccination coverage levels necessary for malaria control under such circumstances. The rest of the paper is organised as follows: In section 2, we present the formulated model together with a flowchart that captures how the different state variables in the model link-up with each other. We further present the theoretical analysis of the mathematical equations in Section 3, model’s parameterization and simulation in Section 4 where we also show results from fitting the model to data, as well as local and global sensitivity analyses. We then round-up the paper with a discussion and conclusions in section 5. Detailed analytical results are presented as supplementary information (SI).

## 2. Model formulation

To study the human-mosquito-ITN-immunity interaction dynamics, we define the following mutually exclusive state variables for *t* ≥ 0: *S*_*h*_(*t*), susceptible unvaccinated humans; *V*_*h*_(*t*), susceptible vaccinated humans; *I*_*h*_(*t*), infectious humans; *R*_*h*_(*t*), recovered (partially immune) humans; *M* (*t*), community immunity level; *E*(*t*), ITN-efficacy; *C*(*t*), ITN-coverage; *S*_*v*_(*t*), susceptible mosquitoes; and *I*_*v*_(*t*), infected mosquitoes. The total human and mosquito populations are *N*_*h*_(*t*) = *S*_*h*_(*t*)+*V*_*h*_(*t*)+*I*_*h*_(*t*)+*R*_*h*_(*t*) and *N*_*v*_(*t*) = *S*_*v*_(*t*)+*I*_*v*_(*t*). Let *λ*_*vh*_ = *λ*_*vh*_(*E, C*) (*λ*_*hv*_ = *λ*_*hv*_(*E, C*)) be the force of infection from mosquitoes (humans) to humans (mosquitoes), then as in [14, 18], 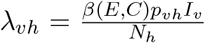 and 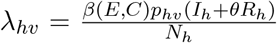, where *p*_*vh*_ (*p*_*hv*_) is the infectivity of infectious mosquitoes (humans) to susceptible humans (mosquitoes), 0 ≤ *θ* ≤ 1 is a modification factor for the infectiousness of recovered humans compared to infectious humans. The biting rate of mosquitoes (*β*(*E, C*)), defined in 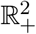 and bounded above by *β*_*max*_ > 0, is modeled with a non-increasing function of both bed net efficacy (*E*) and coverage (*C*).

The susceptible human population (*S*_*h*_) increases through recruitment *f*_*h*_(*N*_*h*_) (assumed to be logistic, linear, or constant), waning of vaccine- and recovery-induced immunity from *V*_*h*_ and *R*_*h*_ at rates *ω*_*p*_ and *ω*_*r*_, respectively, and recovery without immunity at rate *ρ*_*h*_. The susceptible population decreases through vaccination at rate *ξ*_*h*_, natural mortality at rate *µ*_*h*_, and malaria infection at rate *λ*_*vh*_. Thus, the rate of change of *S*_*h*_ is given by the equation: 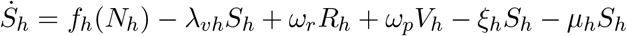, where the dot on *S*_*h*_ denotes differentiation with respect to time (*t*). The population of the vaccinated class (*V*_*h*_) increases through newly vaccinated susceptible individuals (*ξ*_*h*_*S*_*h*_) and decreases due to natural deaths, waning of vaccine-induced immunity at rate *ω*_*p*_, and infection at rate (1 −*ε*_*p*_)*λ*_*vh*_, where 0 ≤ *ε*_*p*_ ≤ 1 is the efficacy of the vaccine that accounts for the reduced susceptibility of individuals in *V*_*h*_ as compared to those in *S*_*h*_. Therefore, the dynamics of *V*_*h*_ is governed by the equation: 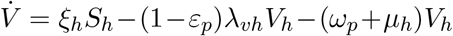. The infectious human class is populated by new infections of susceptible (*λ*_*vh*_*S*_*h*_) and vaccinated ((1 −*ε*_*p*_)*λ*_*vh*_*V*_*h*_) humans and reduce due to natural deaths, disease-related deaths at rate *δ*_*h*_(*M*), recovery without immunity at rate *ρ*_*h*_, and with partial immunity at rate *γ*_*h*_(*M*). It is assumed that *γ*_*h*_(*M*) is a non-decreasing function of *M*, while *δ*_*h*_(*M*) is a non-increasing function of *M*, to highlight the fact that immunity loss leads to severe disease, reducing recovery chances and increasing the death rate. The rate of change of *I*_*h*_ is *İ*_*h*_ = *λ*_*vh*_*S*_*h*_+(1 −*ε*_*p*_)*λ*_*vh*_*V*_*h*_− (*γ*_*h*_(*M*)+*δ*_*h*_(*M*)+*µ*_*h*_)*I*_*h*_. Recovered humans with partial immunity (*γ*_*h*_(*M*)*I*_*h*_) populate the *R*_*h*_ compartment, while the population of the class decreases due to natural mortality and natural immunity waning at a rate *ω*_*r*_. Hence, the rate of change of *R*_*h*_ is 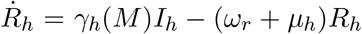. Susceptible mosquitoes (*S*_*v*_) increase through recruitment at rate *f*_*v*_(*N*_*v*_) (assumed logistic or Maynard-Smith-Slatkin with growth rate *r*_*v*_ and carrying capacity *K*_*v*_) and decrease via infection at rate (*λ*_*hv*_) and natural mortality at rate *µ*_*v*_, while infected mosquitoes (*I*_*v*_) increase through this infection process and decrease through natural mortality at rate *µ*_*v*_. Thus, the dynamics of the susceptible mosquito population is governed by the equation 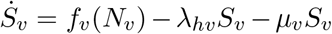, while that of the infectious mosquito population is characterized by *İ*_*v*_ = *λ*_*hv*_*S*_*v*_ − *µ*_*v*_*I*_*v*_.

Population-level immunity is built through exposure 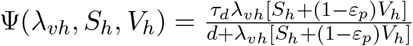, and vaccination, Γ(*V*_*h*_) = *τ*_*p*_*ε*_*p*_*V*_*h*_. Here, *τ*_*d*_ and *τ*_*p*_ are the respective natural and vaccine-derived immunity build-up rates and *d* is the half-saturation constant. Population level immunity decays at rate 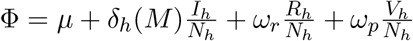, due to human mortality, waning of vaccine immunity, and waning of natural immunity. Hence, the dynamics of population level immunity (*M*) is given by 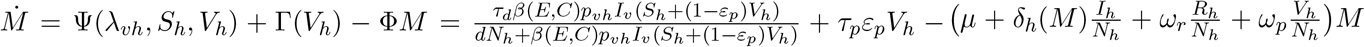. It is assumed that bed net efficacy is restored through replacement at rate *ε*_*e*_ and wanes at rate *σ*_*e*_. The dynamics of bed net efficacy is governed by the equation *Ė* = *ε*_*e*_(*E*_0_ − *E*) − *σ*_*e*_*E*, where *E*_0_ ∈ (0, 1] is the maximum efficacy of bed nets. It is assumed that the rate at which humans use bed nets depends on both the efficacy of the net and the number of mosquitoes visiting human habitats. Based on this, the ITN uptake rate is modeled with the function 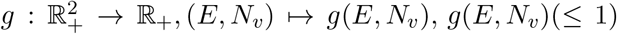, which is a non-decreasing function of both *E* and *N*_*v*_, to capture the fact that ITN-use increases with both ITN-efficacy (*E*) and mosquito abundance (*N*_*v*_), as higher mosquito activity and more effective ITNs encourage greater usage. Moreover, we assume *g*(*E*, 0) = 0, since if there are no mosquitoes in human habitats, then people will not be motivated to use bed nets. Furthermore, it is assumed that bed net coverage decreases at a rate *σ*_*c*_. Thus, the bed net coverage equation is *Ċ* = *g*(*E, N*_*v*_)(*C*_0_ − *C*) − *σ*_*c*_*C*, where *C*_0_ ∈ [0, 1] is the maximum coverage rate.

Combining the equations governing the dynamics of susceptible, vaccinated, infectious, and recovered humans, population immunity, ITN efficacy and coverage, and the susceptible and infectious mosquito populations, together with the parameter definitions and flow diagram in Table 1 and Figure 1, respectively, yields System (2.1).

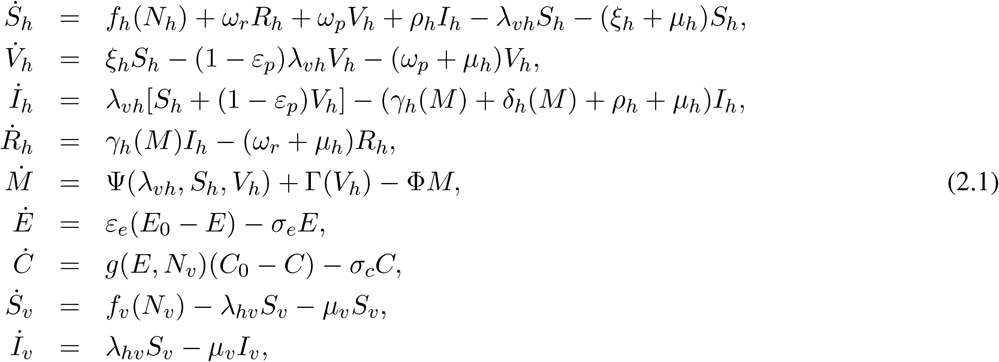

where 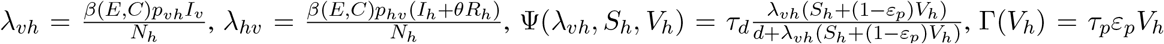, and 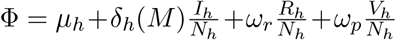. The dynamics of the total human and mosquito populations are described by the equations 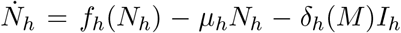 and 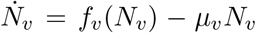, while the model system (2.1) is subject to the initial conditions *S*_*h*_(0) = *Sh*_0_ > 0, *V*_*h*_(0) = *V*_*h*0_ ≥ 0, *I*_*h*_(0) = *I*_*h*0_ ≥ 0, *R*_*h*_(0) = *R*_*h*0_ ≥ 0, *S*_*v*_(0) = *S*_*v*0_ > 0, *I*_*v*_(0) = *I*_*v*0_ ≥ 0, *M* (0) = *M*_0_ > 0, *E*(0) = *E*_0_ ≥ 0, *C*(0) = *C*_0_ ≥ 0.

**Table 1.** Parameters of the model (2.1) and their epidemiological interpretations and units.

| Parameter | Epidemiological interpretation | Unit |
| --- | --- | --- |
| $\beta$ | Mosquitoes biting rate. | Bites per year |
| $p_{vh}$ | Infectivity of mosquitoes to humans. | Human per bite per mosquito. |
| $p_{hv}$ | Infectivity of humans to mosquitoes. | Per bite |
| $\theta$ | Modification parameter for the infectiousness of recovered humans. | 1 |
| $\xi_h$ | Vaccination rate. | per year |
| $\varepsilon_p$ | Vaccine efficacy. | 1 |
| $\omega_p(\omega_r)$ | Immunity waning rate in $V_h(R_h)$ compartment. | per year |
| $\rho_h$ | Malaria recovery rate without immunity. | per year |
| $\mu_h(\mu_v)$ | Natural mortality rate of humans (mosquitoes). | per year |
| $\gamma_h$ | Malaria recovery rate. | per year |
| $\delta_h$ | Malaria-induced death rate. | per year |
| $\tau_p$ | Immunity rate acquired by vaccination. | Immunity per human per year |
| $\tau_d$ | Immunity build-up rate per mosquito bite. | Immunity per year |
| $d$ | Half saturation constant. | Humans per year |
| $\sigma_c$ | Decay rate of bed net coverage. | per year |
| $\sigma_e$ | Decay rate of bed net efficacy. | per year |
| $\varepsilon_e$ | Bed net replacement rate. | per year |

**Figure 1.**
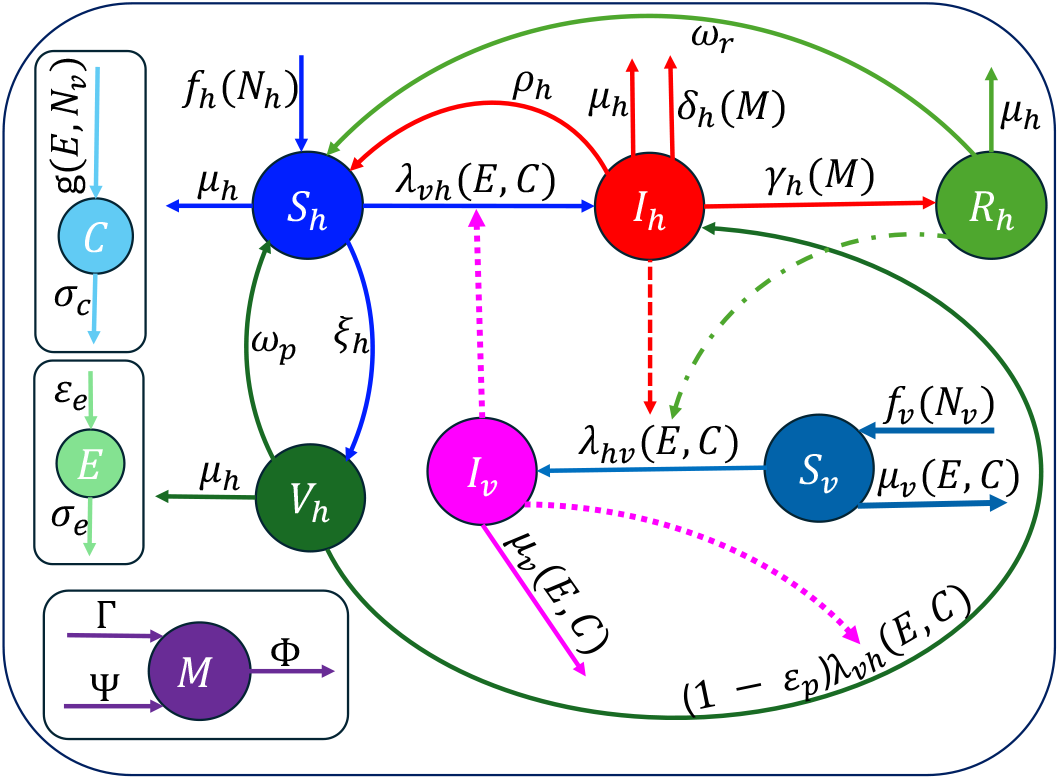
Flow diagram of Model (2.1). A conceptualisation of the human-mosquito-bed net-immunity dynamic epidemiological model. Human compartments, identified with the *h*-subscript, follow an Susceptible-Vaccinated-Infectious-Recovered (*SV IR*) compartmental model, the mosquito subclasses, identified with the *v*-subscript, follow the *SI* compartmental model. The total human and mosquito populations are denoted by *N*_*h*_ and *N*_*v*_, respectively. Population-level immunity is represented by the compartment *M*, accumulates through infection-driven processes Ψ = Ψ(*λ*_*vh*_, *S*_*h*_, *V*_*h*_) and vaccination Γ = Γ(*V*_*h*_), and wanes at rate Φ. The effects of insecticide-treated nets are captured by the efficacy and coverage compartments *E* and *C*. Transmission from infectious mosquitoes to susceptible and vaccinated humans is illustrated by dotted lines, while transmission from infectious and recovered humans to susceptible mosquitoes is represented by dashed and dash-dotted lines, respectively. The dynamics within and between the compartments, as well as the transition rates are as explained in the text.

## 3. Theoretical analysis of Model (2.1)

In this section, the model (2.1) is studied analytically for the case with general human and mosquito recruitment terms (*f*_*h*_ and *f*_*v*_) and for various representative combinations of *f*_*h*_ and *f*_*v*_ (summarized in Table S1 of the SI) to gain insights into the influence of recruitment function on the dynamics of the model. Specific functional forms for *f*_*h*_ and *f*_*v*_ include the constant, linear, logistic, and Maynard-Smith-Slatkin functions. Since all state variables of the model (2.1) are non-negative as they represent populations and approximately measurable quantities, it can be verified that the model system (2.1) is well-posed, admitting a unique global solution that remains nonnegative, evolves within a positively invariant bounded region (Ω), and whose trajectories ultimately enter and remain in this biologically feasible domain. This is formalized in Theorem 3.1, whose proof is provided in Section S1.1 of the SI.

### Theorem 3.1.

*The model* (2.1) *defines a dynamical system on the biologically feasible compact region* 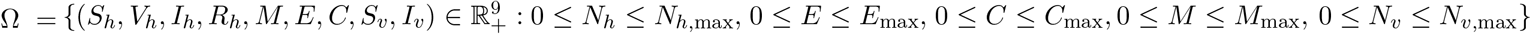, *which is positively invariant and attracting*.

### 3.1 Analysis of the model (2.1) for various combinations of human and mosquito recruitment functions

For epidemiological realism, the transmission, recovery, and disease-induced death rates are allowed to vary dynamically in response to key epidemiological and control drivers, thereby capturing nonlinear feedback effects induced by intervention coverage, immunity development, and vector control pressure. Specifically, 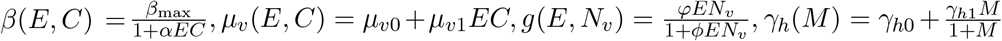, and 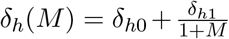. In this formulation, *β*(*E, C*) decreases with increasing combined ITN effectiveness *EC*, reflecting reduced mosquito–human contact under stronger protection. The mosquito mortality rate *µ*_*v*_(*E, C*) increases with ITN coverage due to intervention-induced vector mortality. The uptake of ITNs is modeled by the saturating function *g*(*E, N*_*v*_), which increases with both effort *E* and mosquito population *N*_*v*_ but saturates at high deployment levels due to logistical and behavioural constraints. The epidemiological functions, *γ*_*h*_(*M*) and *δ*_*h*_(*M*), capture immunity-dependent recovery and disease-induced mortality, respectively. The recovery rate *γ*_*h*_(*M*) increases with immunity but saturates, reflecting diminishing returns at high immune levels, while *δ*_*h*_(*M*) decreases with increasing immunity burden effects, consistent with reduced severity or improved host tolerance at higher immune states. Here, *β*_max_ is the maximum mosquito biting rate in the absence of ITNs, *α* measures the efficacy of bed nets in reducing effective contact, *µ*_*v*0_ is the baseline mosquito mortality rate, and *µ*_*v*1_ captures ITN-induced mosquito mortality. The parameters *φ* and *ϕ* govern the intensity and saturation of ITN uptake, while *γ*_*h*0_, *γ*_*h*1_, *δ*_*h*0_, and *δ*_*h*1_ define baseline and immunity-regulated recovery and mortality dynamics. In particular, *δ*_*h*0_ and *γ*_*h*0_ are the baseline malaria-induced death and recovery rates, while 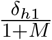 and 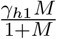 measure the influence of immunity on malaria death and recovery.

#### 3.1.1 Existence and stability of disease-free equilibria and the reproduction number of the model

Disease-free equilibria, 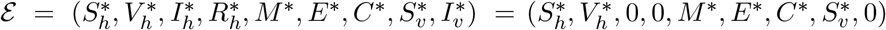, of the model (2.1) are obtained by setting *I*_*h*_ = *I*_*v*_ = 0, which forces *λ*_*vh*_, *λ*_*hv*_ and *R*_*h*_ = 0 to be zero, and then equating the left-hand sides of the model equations to zero. Solving the resulting system of equations simultaneously, leads to up to three possible disease-free equilibria that can be expressed explicitly in closed form: 1) An extinction equilibrium 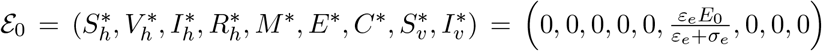, always exists for any choice of *f*_*h*_ and *f*_*v*_ provided that *f*_*h*_(0) = 0 and *f*_*v*_(0) = 0. 2) A mosquito-free equilibrium 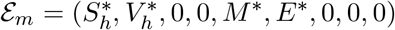, where 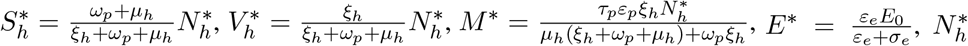 is the unique positive solution of the equation 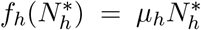. 3) The full disease-free equilibrium 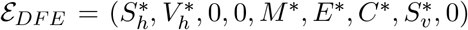, where 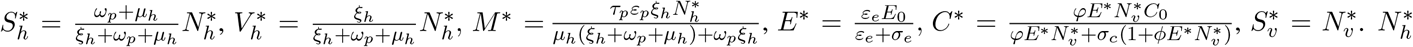 is the positive solution to the host demographic equation 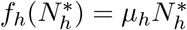, and 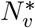 is the positive solution of the equation, 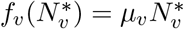.

The reproduction number of the model (2.1) obtained through the next generation operator approach is

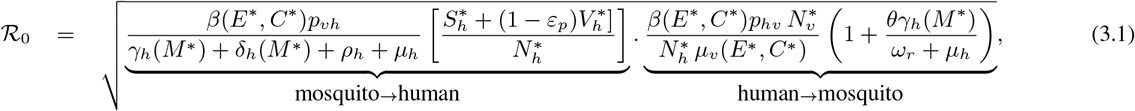

where 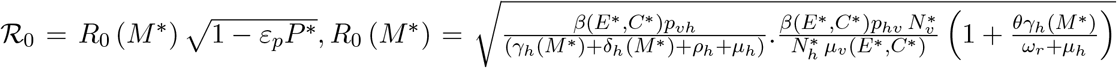, with 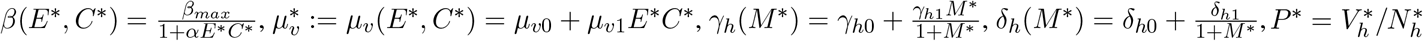 is vaccine coverage. Since 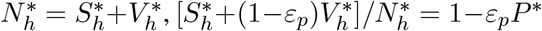. The reproduction number (ℛ_0_) captures malaria transmission as a multiplicative interplay of vaccination, vector control, and immunity dynamics. Vaccination reduces susceptibility via 1 *ε*_*p*_*P* ^∗^, ITNs suppress transmission through (1 + *αE*^∗^*C*^∗^)^−2^, and vector control increases mortality via *µ*_*v*0_ + *µ*_*v*1_*E*^∗^*C*^∗^. These effects are reinforced by immunity-dependent processes through *γ*_*h*_(*M* ^∗^) and *δ*_*h*_(*M* ^∗^), which shorten infectious duration alongside *ρ*_*h*_ + *µ*_*h*_. Thus, ℛ_0_ reflects a synergistic multi-layer control system in which vaccination limits susceptibility, ITNs suppress transmission and vector longevity, and immunity processes constrain infectious persistence, together determining whether transmission can be sustained or contained. Epidemiologically, ℛ_0_ represents the expected number of secondary infections generated by a single infectious individual in a structured population where vaccination reduces effective susceptibility, while ITNs reduce transmission and increase vector mortality. From a public health perspective, ℛ_0_ < 1 guarantees local stability of the disease-free equilibrium and disease elimination, whereas ℛ_0_ > 1 implies invasion and persistence; increasing vaccine uptake or efficacy (*ε*_*p*_) reduces ℛ_0_ and therefore lowers outbreak risk. Hence, we have the following results, whose proofs are presented in Section S1.2 of the SI.

##### Theorem 3.2.

*The model* (2.1) *admits up to three disease-free equilibria: (1) an extinction equilibrium, which is locally asymptotically stable when* 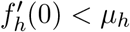 *and* 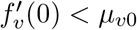, *(2) a mosquito-free equilibrium, which is locally asymptotically stable when* 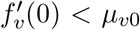, *and (3) a full disease-free equilibrium with vector persistence, which is locally asymptotically stable if* ℛ_0_ < 1 *and unstable if* ℛ_0_ > 1.

##### Theorem 3.3.

*(i) The extinction equilibrium of the model* (2.1) *is globally asymptotically stable when* 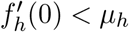 *and* 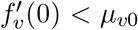. *(ii) The mosquito-free equilibrium of Eqs*. (2.1) *is globally asymptotically stable when* 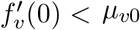. *(iii) The full disease-free equilibrium of Eqs*. (2.1) *is globally asymptotically stable when* ℛ_0_ ≤ 1.

#### 3.1.2 Analysis of the model (2.1) with constant human and Maynard-Smith-Slatkin mosquito recruitment

As an illustrative example, we analyze the full model (2.1) under constant human recruitment (*f*_*h*_(*N*_*h*_) = Λ_*h*_) and Maynard-Smith-Slatkin mosquito recruitment 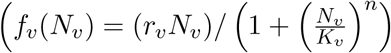, combining fixed host inflow with density-dependent vector regulation. This formulation provides a biologically realistic framework for studying how recruitment structure shapes equilibrium existence and disease dynamics. Notably, as shown in [21, 32, 33], the Maynard-Smith-Slatkin function is more appropriate than logistic growth for vector dynamics, as it ensures mosquito persistence in the ecosystem and leads to rich dynamical behaviours.

The mosquito-free equilibrium (MFE) of the full model (2.1) is given by 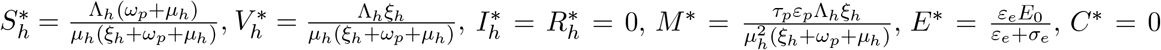, and 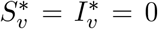, while the disease-free equilibrium (DFE) is given by 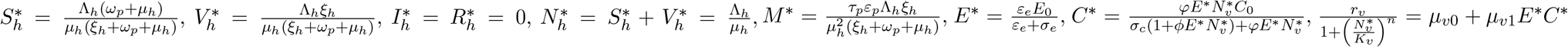, and 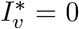. Since *C*^∗^ and 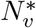 are mutually dependent, with each defined in terms of the other, substituting the equilibrium expression of *C*^∗^ into the equation for 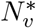 and simplifying yields the polynomial equation 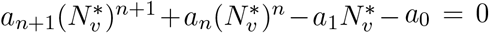, where the explicit coefficients are: 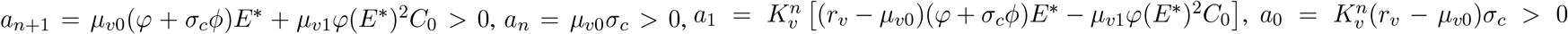, since *r*_*v*_ *> µ*_*v*0_. Since *a*_*n*+1_, *a*_*n*_, *a*_0_ > 0, irrespective of whether *a*_1_ is positive or negative, there is exactly one sign change in the sequence of coefficients. Therefore, according to Descartes’ Rule of Signs, the equation has at most one possible positive real solution 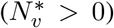. Thus, the model has a single, well-defined, and biologically meaningful non-trivial disease-free equilibrium. Details of the calculations are presented in Sections S1.2-S1.4 and S2 of the SI.

For this case of constant human and Maynard-Smith-Slatkin mosquito recruitment, the vaccination-ITN-controlled reproduction number in (3.1) reduces to 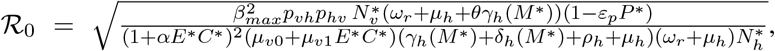 where 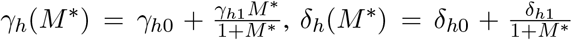. Details of the calculations and other examples with other recruitment functional forms are presented in Sections S1.3 and S2 of the SI.

## 4.Model parameterization and simulation

### 4.1. Model parameterization

Some parameters of Model (2.1) were obtained directly from malaria epidemiological data and demographic characteristics of human and mosquito populations, others were sourced from published literature (Table 2 (a)), and the remainder (Table 2 (b)) are estimated by fitting the model to available confirmed malaria data from Kenya [34]. Specifically, fixed model parameters were sourced from the literature or inferred from empirical epidemiological and demographic data to ensure biologically realistic and context-specific calibration.

**Table 2.** Fixed and estimated parameter values for the full malaria transmission model (2.1), with dynamic insecticide-treated net (ITN) use, vaccination, constant human recruitment, and Maynard-Smith-Slatkin mosquito recruitment. (a) Summary of fixed parameters used for simulation and calibration together with plausible ranges and references. (b) Estimated parameter values obtained using nonlinear least-squares fitting with bootstrap-based uncertainty quantification (1000 bootstrap samples). Estimates are accompanied by 95% confidence intervals (CI). The estimated reproduction number was ℛ_0_ = 1.4119 (95% CI: [1.3424, 1.4832]). All rates are expressed in units of *per year*, the human recruitment rate (Λ_*h*_) in *humans per year*, and the mosquito carrying capacity (*K*_*v*_) in mosquitoes, while *n*, all probabilities, and proportions are dimensionless.

| (a) Fixed parameter values and their ranges. |  |  |  | (b) Estimated parameter values and their 95% CIs. |  |  |
| --- | --- | --- | --- | --- | --- | --- |
| Parameter | Value | Range | Source | Parameter | Value | 95% CI |
| $\theta$ | 0.33 | [0.072, 0.640] | [44] | $\eta$ | 0.38780 | [0.30529, 0.52605] |
| $\mu_h$ | 1/65 | [1/70, 1/60] | [36] | $\tau_d$ | 6.65920 | [0.72865, 9.44500] |
| $\mu_{v0}, \mu_{v1}$ | 365/21 | [365/30, 365/14] | [44] | $p_{vh}$ | 0.18702 | [0.17490, 0.20372] |
| $\delta_{h0}$ | 0.0329 | [0, 0.15] | [44] | $\delta_{h1}$ | 0.54737 | [0.07421, 0.93934] |
| $p_{hv}$ | 0.3735 | [0.072, 0.64] | [44] | $\varphi$ | 1.28970 | [1.05860, 2.58070] |
| $\gamma_{h0}, \rho_h$ | 365/14 | [365/21, 365/7] | [38–41] | $\phi$ | 0.45867 | [0.34101, 0.84486] |
| $\omega_r$ | 1/12 | [1/16, 1] | [42–44] | $n$ | 2.25020 | [2.00140, 2.95720] |
| $E_0$ | 0.65 | [0.40, 0.70] | [50, 51] | $\gamma_{h1}$ | 7.40060 | [4.0188, 12.94700] |
| $C_0$ | 0.90 | [0.85, 0.95] | [50–52] | $\alpha$ | 9.71590 | [8.53100, 11.0910] |
| $\omega_p$ | 0.40 | [0.20, 0.60] | [57, 58] | $\tau_p$ | 6.79160 | [0.83766, 9.42660] |
| $\varepsilon_e$ | 0.32 | [0.25, 0.40] | [54, 55] | $d$ | 409.150 | [47.6210, 484.370] |
| $\sigma_e$ | 0.27 | [0.20, 0.35] | [53] | $\beta_{\max}$ | 46.6660 | [45.1070, 49.9740] |
| $\varepsilon_p$ | 0.35 | [0.20, 0.50] | [59] | $r_v$ | 29.3890 | [25.4450, 31.1740] |
| $\sigma_c$ | 0.15 | [0.05, 0.45] | [52, 56] | $K_v$ | $5.5426 \times 10^8$ | $[4.1375, 7.3799] \times 10^8$ |
| $\xi_h$ | 0.31 | [0.27, 0.37] | [59] | $\Lambda_h$ | $1.7592 \times 10^6$ | $[1.5891, 2.3720] \times 10^6$ |

The human recruitment rate was estimated by fitting Kenyan population data from Ref. [35] to a population model as in [26], yielding Λ_*h*_ = 1,759,200 persons per year. The natural human mortality rate is set to *µ*_*h*_ = 1/65 *per year* with a range of [1/74, 1/56] *per year*, based on an average life expectancy of 65 [56, 74] years [36]. The malaria-induced mortality rate was set to *δ*_*h*0_ = 32.9 ([0.00, 150]) × 10^−3^ *per year* [37], corresponding to a case-fatality ratio of approximately 0.13 % per infection, which is consistent with low-to-moderate malaria mortality settings such as Kenya. *P. falciparum* infections are acute and rapidly cleared with treatment but may persist for months if untreated or asymptomatic. Based on a study that reported untreated durations of up to 21 days [38–41], we set *γ*_*h*0_ = 365/14 ([365/21, 365/7]) *per year*) and a treatment duration of 14 ([7, 21]) days, which results in *ρ*_*h*_ = 365/14 ([365/21, 365/7]) *per year* [38–41]. Naturally acquired immunity to *P. falciparum* is partial and non-sterilizing, sustained by re-exposure but waning without it. We assume a baseline duration of 12 ([1, 16]) years, i.e., *ω*_*r*_ = 1/12 ([1/16, 1]) *per year*, consistent with evidence of long-lived immune memory and shorter-lived functional protection from Ref. [42–44]. To align with the findings of Idris *et al*. [45], the infectiousness modification parameter for recovered individuals is set to *θ* ≈ 0.33 ([0.072, 0.640]). This adjustment reflects the observation that the parasite density and the level of infectiousness of asymptomatic (recovered) individuals is approximately one-third [46] that of symptomatic (actively infectious) individuals. Adult malaria vectors (*Anopheles* spp.) typically live about 2–4 weeks, with field estimates often around 10-14 days and rarely exceeding one month [47]. Using a baseline lifespan of 21 days gives a natural mortality rate *µ*_*v*0_ = 365/21 ([365/30, 365/14]) *per year*. The maximum ITN-induced mosquito mortality rate was set to *µ*_*v*1_ = 365/21 ([365/30, 365/14]) *per year*. This is supported by experimental hut and field studies showing substantial, repeated-contact mortality from treated nets. [48, 49]. As in [44], the human to mosquito transmission probability is set to *p*_*hv*_ = 0.373 ([0.072, 0.640]).

Parameters related to ITN-use are derived from Kenyan field studies information. In particular, the maximum efficacy of ITNs is set to *E*_0_ = 0.65 with a range of [0.40, 0.70], which is consistent with individual-level infection reductions of 23%-43% and population-level reductions up to ~ 70% [50, 51]. The maximum ITN coverage (*C*_0_) is in the range *C*_0_ ∈ [0.85, 0.95] with an average value of *C*_0_ = 0.9 [50–52]. The decay rate of ITN-efficacy is in the range *σ*_*e*_ ∈ [0.20, 0.35] *per year* (average *σ*_*e*_ ≈ 0.27) [53], corresponding to an approximately 2.9-5 (3.7) years ITN lifespan, while ITN replacement occurs at rate *ε*_*e*_ ≈ 0.32 ([0.25, 0.40]) *per year* [54, 55]. The abandonment rate of ITN-usage is *σ*_*c*_ ≈ 0.15 ([0.05, 0.45]) *per year* [52, 56]. Parameterization of the malaria vaccine component is based on clinical trials and longitudinal field data. For RTS,S/AS01, efficacy is moderate, *ε*_*p*_ ∈ [0.30, 0.40] (mean ≈ 0.35), with protection waning within 2-3 years, corresponding to *ω*_*p*_ ∈ [0.33, 0.50] *per year* (average, ≈ 0.4) [57, 58]. The vaccination rate is *ξ*_*h*_ ∈ [0.27, 0.37] *per year* (mean ≈ 0.31), derived from the four-dose completion range, [0.40, 0.55] over an ≈ 1.5-year schedule [59]. In contrast, the vaccine (R21/Matrix-M) has a higher short-term efficacy, *ε*_*p*_ ∈ [0.66, 0.75] (mean ≈ 0.70), sustained for ≈ 12 months following a three-dose series [60]. Its vaccination rate is *ξ*_*h*_ ∈ [0.70, 0.85] *per year* (mean ≈ 0.78 *per year*), with waning *ω*_*p*_ ∈ [0.08, 0.15] *per year* (mean ≈ 0.11), reflecting slower decay and booster-supported durability [60].

Estimation of the remaining parameter values was carried out by fitting two versions of the mechanistic malaria transmission model (2.1)–the full model and a reduced version of the model with no vaccination, to reported annual malaria incidence data for Kenya that spans the period from 2002 to 2022. The reduced model was considered because malaria vaccination was introduced only recently and therefore had limited influence over much of the fitting period. The dataset comprises 20 consecutive yearly incidence observations (cases per year), beginning with ≈ 20, 000 reported cases in the first year and ending with ≈ 4.9 million cases in the last year of the study period. For consistency between the reported data and the model outputs, we introduced a virtual detection (or confirmation) compartment with reporting rate *η* that represents diagnosed individuals reported through the health system. The model parameter values were estimated using nonlinear least squares, a statistical optimization technique that minimizes the sum of the squared differences between the observed malaria incidence data and the incidence values derived from the model. This objective function ensures that the resulting parameter set yields a trajectory that most closely aligns with the empirical data. Minimization of the objective function was performed using MATLAB’s `lsqcurvefit` function. The model fit is depicted in Fig. 2(a) and (d), and the estimated parameter values for the full model are presented in Table 2 (b), while those for the reduced model are presented in Table S2 of the SI. An illustration of the long-term dynamics of the model beyond the fit time window is shown in Figs. S3-S4 of the SI.

**Figure 2.**
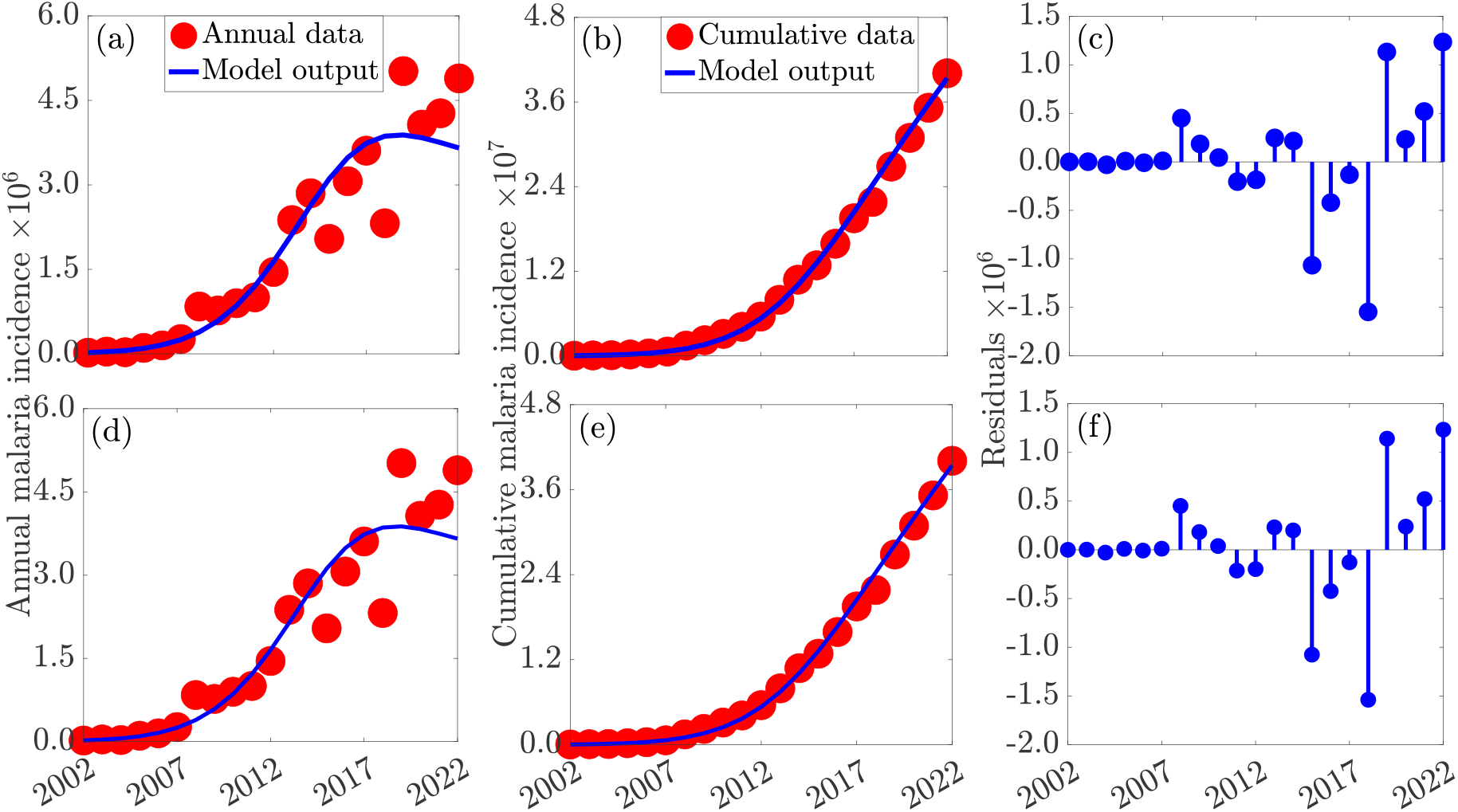
Calibration and validation of the full malaria transmission model (2.1) ((a)-(c)) and the model (2.1) with no vaccination ((d)-(f)) using confirmed annual malaria incidence data from Kenya. (a) and (d): A comparison of the model fit (solid blue curve) against observed annual confirmed malaria cases (filled red circles) illustrating the ability of the calibrated model to reproduce the long-term temporal patterns in reported incidence. (b) and (e): Model validation by comparing cumulative confirmed cases generated using the estimated parameter values together with the fixed parameter values reported in Tables 2 against the corresponding cumulative observed data, providing an assessment of the model’s ability to capture the aggregate disease burden over time beyond year-to-year fluctuations. (c) and (f): Residuals, highlighting the magnitude and temporal structure of discrepancies between model outputs and surveillance data and providing a visual assessment of goodness-of-fit, systematic bias, and remaining unexplained variability.

To quantify parameter uncertainty, we used a parametric residual bootstrap approach with 1000 replications. Empirical residuals from the primary deterministic fit were centered to preserve unbiasedness and resampled with replacement to generate synthetic data trajectories. The model was then re-fitted to each of these 1000 bootstrap replicas, generating a distribution of parameter estimates from which the 95% confidence intervals were calculated. To compute the 95% confidence interval of the reproduction number (ℛ_0_), ℛ_0_ was calculated at the disease-free equilibrium for each individual bootstrap iteration. This process translated the underlying parameter variations and interactions directly into the 95% confidence interval of the derived ℛ_0_. The 95% confidence are displayed in Table 2 (b). A collection of profile histograms representing the empirical bootstrap distributions for the estimated model parameters (blue histograms in Fig.S1 (a)-(n) of the SI) and the derived ℛ_0_ (green histogram in Fig. S1 (o)-S2 (n) of the SI) is presented in Fig. S1-S2 of the SI. For each subplot, the horizontal axis represents the estimated numerical range of the specific parameter or ℛ_0_, while the vertical axis represents the frequency count of the refitted values across all bootstrap iterations. The results are interpreted by assessing the width of the histogram and the distance between the dotted vertical magenta lines, which define the 95% confidence interval and indicate parameter certainty. A tight clustering around the solid red line (the original point estimate) signifies a highly identifiable, unbiased parameter, whereas the blue ℛ_0_ histogram statistically confirms sustained transmission potential if its entire distribution sits strictly above the threshold value of ℛ_0_ = 1 as is the case here.

To assess the model’s performance, we analyzed statistical indicators alongside a direct simulation comparison. The model achieved a coefficient of determination of *R*^2^ = 0.8755, implying that it successfully explains roughly 88% of the variance in the observed data. We then used the estimated and fixed parameter set in Table 2 to simulate the model, verifying that the cumulative case curve from the model mirrors the aggregated data closely (Fig. 2(b)). Furthermore, residual analysis showed that deviations between the observed and predicted values fluctuate randomly around zero without systematic patterns (Fig. 2(c)). The initial conditions used are: *S*_*h*0_ = 32, 609, 759, *V*_*h*0_ = 0, *I*_*h*0_ = 55, *R*_*h*0_ = 100, 000, *M*_0_ = 150, *E*_0_ = 0.65, *C*_0_ = 0.1, *S*_*v*0_ = 163, 139, 375, and *I*_*v*0_ = 9, 665, where *I*_*h*0_ and *I*_*v*0_ are chosen such that the initial incidence from the model matches the first incidence data point.

### 4.2. Simulations

Since both the full model and the reduced model without vaccination provide comparable fits to the observed incidence data, we carry out subsequent simulations with a the full model. From a public health perspective, a good fit to historical data alone is not sufficient for evaluating future control strategies. The full model incorporates vaccination explicitly, a key intervention that is increasingly being integrated into malaria control programs and is expected to play an important role in reducing transmission and disease burden. Retaining the vaccination component therefore allows the model to capture both current transmission dynamics and the potential impact of future vaccination scale-up. Hence, in this section, unless otherwise stated, the model (2.1) is simulated using the initial conditions *S*_*h*0_ = 32, 609, 759, *V*_*h*0_ = 0, *I*_*h*0_ = 55, *R*_*h*0_ = 100, 000, *M*_0_ = 150, *E*_0_ = 0.65, *C*_0_ = 0.1, *S*_*v*0_ = 163, 139, 375, and *I*_*v*0_ = 9, 665, together with the parameter values provided in Tables 2, to assess various epidemiological scenarios including different intervention and transmission settings.

#### 4.2.1. Long-term dynamics and backward bifurcation

To gain insight into the relationship between the basic reproduction number, ℛ_0_, and the long-term behaviour of the system (2.1), we investigated model dynamics in a small neighborhood of the threshold value ℛ_0_ = 1 numerically. The results shown in Fig. 3(a)-(d) indicate that, for the baseline parameter set, ℛ_0_ behaves as the expected epidemiological threshold. When ℛ_0_ = 0.999, solutions converge to the disease-free equilibrium, whereas a slight increase to ℛ_0_ = 1.001 leads to convergence toward a stable endemic state. Thus, even an extremely small perturbation across the threshold separates eventual disease elimination from long-term persistence. For clarity, only a zoomed-in portion of the dynamics is shown here; the corresponding full trajectories are presented in Fig. S5 of the SI. To explore whether this threshold behaviour remains robust under altered epidemiological conditions, we repeated the analysis after increasing the malaria-induced mortality rate to 1000 times its baseline value while leaving all other parameters unchanged. The resulting dynamics, displayed in Fig. 3(e)-(h), reveal a markedly different scenario characterized by the presence of a backward bifurcation (see Fig. S6 of the SI also). In this case, the disease-free equilibrium and a stable endemic equilibrium coexist for a range of parameter values with ℛ_0_ < 1. As a consequence, reducing ℛ_0_ below unity is no longer sufficient to guarantee elimination. Instead, the eventual outcome depends on the initial disease burden, with sufficiently large outbreaks converging to endemic persistence even when the reproduction number is below one. From a public health perspective, this phenomenon is particularly important because it implies that achieving ℛ_0_ < 1 may not, by itself, be enough to eliminate malaria. Additional efforts may be required to reduce prevalence below a critical threshold before elimination becomes attainable. Broadly, these results underscore the need for caution when interpreting ℛ_0_ as the sole measure of control success in settings where nonlinear epidemiological mechanisms can generate bistability and backward bifurcation.

**Figure 3.**
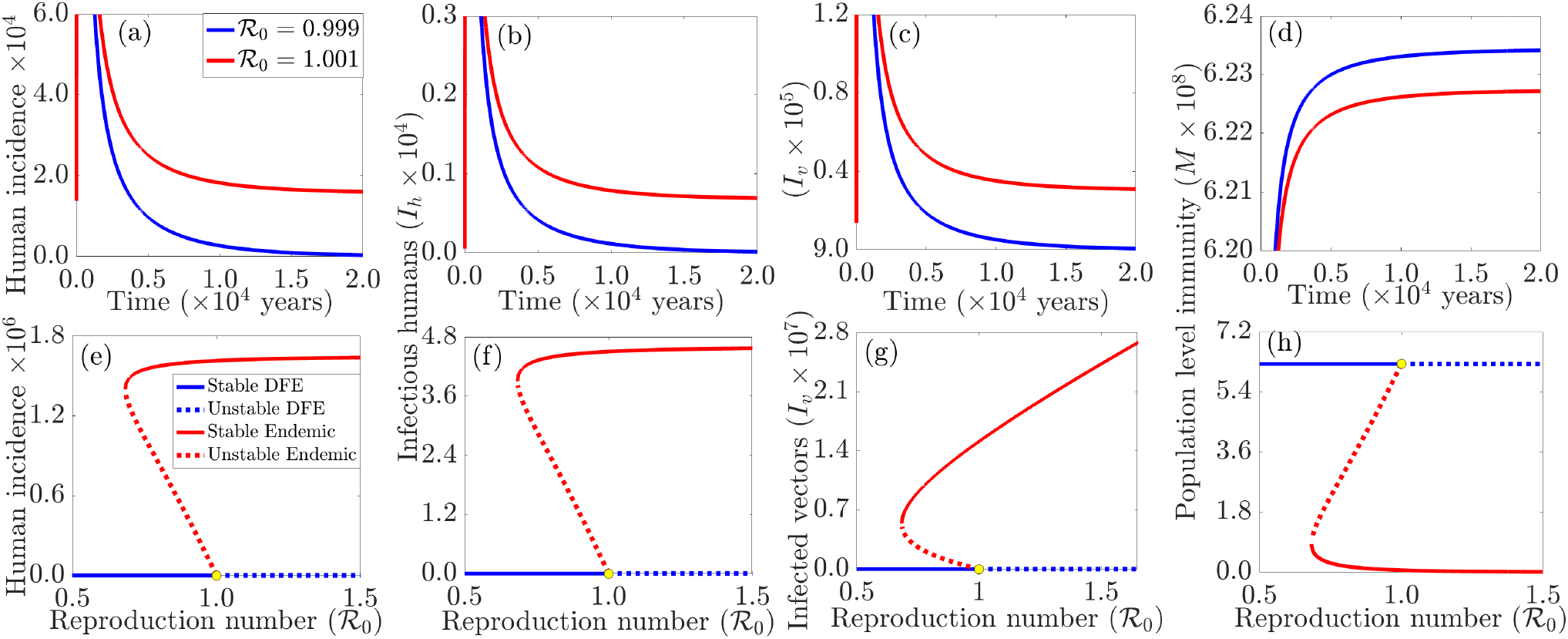
Long-term dynamics of the malaria model illustrating the threshold behaviour of ℛ_0_ ((a)-(d)), and the emergence of backward bifurcation under a high malaria-induced mortality rate ((e)-(h)). Panels (a)-(d) show convergence to the disease-free equilibrium for ℛ_0_ = 0.999 and to an endemic equilibrium for ℛ_0_ = 1.001, confirming the classical threshold property of ℛ_0_. Panels (e)-(h) show the coexistence of stable disease-free and endemic equilibria when the malaria-induced mortality rate is increased to 1000 times its baseline value, demonstrating the occurrence of backward bifurcation. The simulations were initiated in 2002 using the initial conditions *S*_*h*_(0) = 32,609,759, *V*_*h*_(0) = 0, *I*_*h*_(0) = 55, *R*_*h*_(0) = 100,000, *M* (0) = 150, *E*(0) = 0.65, *C*(0) = 0.1, *S*_*v*_(0) = 163,139,375, and *I*_*v*_(0) = 9,665, together with the parameter values listed in Tables 2.

#### 4.2.2. Assessing the impacts of ITN and vaccination on malaria incidence and acquired immunity

Naturally acquired immunity to malaria is boosted by repeated exposure to infectious mosquito bites. However, ITN-use reduces this exposure (since less mosquito biting translates into less exposure to malaria infection), raising an important question about whether the resulting decline in acquired immunity could offset the overall benefits of malaria prevention. To assess whether ITNs are beneficial or detrimental to malaria control, the model (2.1) was simulated under fixed ITN coverage levels of 0%, 25%, 50%, 75%, 90%, and 100%. The 90% coverage corresponds to the baseline parameterization used throughout this study and will serve as a reference point. To isolate the direct effects of ITN coverage, the feedback mechanisms governing changes in ITN-use were disabled, ensuring that coverage remained constant during each simulation. The results shown in Fig. 4(a)-(b) highlight the impact of different ITN coverage levels relative to the baseline coverage of 90%. In the absence of ITNs (0% coverage), equilibrium malaria incidence will be approximately 124% higher than the baseline level, while population-level immunity will be 99% lower. At 25% coverage, malaria incidence is 112% higher than the baseline, whereas population-level immunity is 93% lower. At 50% coverage, incidence is 87% higher and population-level immunity is 72% lower than the baseline values. Increasing coverage to 75% will substantially reduce these differences, although incidence will still be 41% higher and immunity 30% lower than under the baseline coverage scenario. In contrast, increasing coverage from the baseline level of 90% to full (100%) coverage will result in an additional 35% reduction in equilibrium malaria incidence and a 21% increase in immunity. These results show that progressively higher ITN coverage is associated with substantial reductions in malaria burden and improvements in population-level immunity, with the greatest benefits occurring as coverage approaches universal levels. Hence, increasing ITN coverage reduces human exposure to infectious mosquito bites by lowering the frequency of mosquito–human contact, while also directly reducing mosquito populations through increased mosquito mortality and reduced survival and reproduction. This dual effect decreases transmission and leads to a lower force of infection in the human population, which in turn reduces naturally acquired immunity over time due to fewer infectious exposures. However, the decline in transmission is accompanied by a substantially larger reduction in malaria incidence, with clinical cases becoming very low at high coverage levels. Therefore, the combined impact of reduced biting and reduced mosquito population dominates any loss of exposure-induced immunity, making sustained high ITN coverage highly effective for reducing malaria burden and preventing clinical disease at the population level.

**Figure 4.**
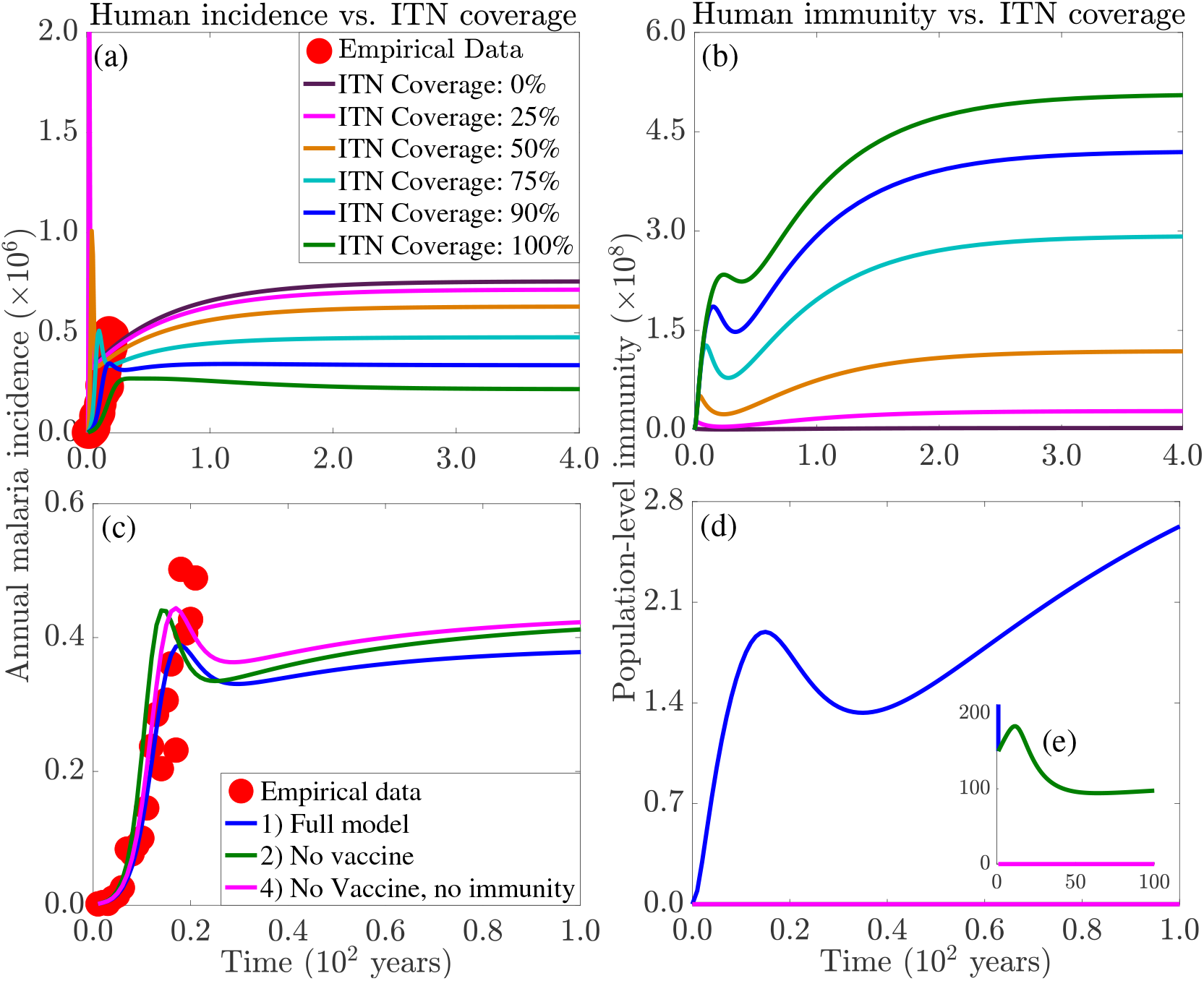
Effect of fixed ITN coverage on long-term malaria dynamics. (a) Human disease incidence and (b) population-level naturally acquired immunity for ITN coverage levels ranging from 0% to 100%. Increasing ITN coverage reduces exposure-driven immunity but produces a substantially larger reduction in malaria incidence, indicating that the transmission-blocking benefits of ITNs outweigh the loss of acquired natural immunity. This reinforces the importance of sustained high ITN coverage for malaria control. (c)–(d): Comparative impact of vaccination on malaria case reduction. The full model with vaccination reduces more cases compared to a model with ITN-use as the only control strategy. The simulations were initiated in 2002 using the initial conditions *S*_*h*_(0) = 32,609,759, *V*_*h*_(0) = 0, *I*_*h*_(0) = 55, *R*_*h*_(0) = 100,000, *M* (0) = 150, *E*(0) = 0.65, *C*(0) = 0.1, *S*_*v*_(0) = 163,139,375, and *I*_*v*_(0) = 9,665, together with the parameter values listed in Tables 2.

Insecticide treated net coverage reduces acquired immunity because less mosquito biting translates into less exposure to malaria and less opportunity to acquire immunity. However, overall immunity trends upward with coverage due to the ITNs modeled in Figure 4(b). This is because immunity is not only acquired through malaria infection but also through vaccination. With lower force of infection due to ITN coverage, more vaccinated people can retain their vaccine protection, hence acquiring immunity from it to a level that overrides the waning immunity acquired from malaria infection. Consequently, the increase in immunity reflects the combined effects of vaccination and reduced disease burden rather than increased malaria transmission.

To determine the extent of population-level immunity and burden reduction due to vaccination, three scenarios spanning the model (2.1) with vaccination, no-vaccination, and no-vaccination/no-immunity (susceptible individuals from non-endemic areas or returning residents of non-endemic areas, exposed to malaria infection for the first time) are compared. As shown in Fig. 4(c)–(d), incorporating vaccination produces a clear reduction in disease burden and an increase in population-level immunity relative to the version of the model with ITN-use only. Specifically, compared with the no-vaccination scenario, the baseline model yields an approximately 13% reduction in peak incidence and a 6% (9%) lower incidence by year 50 (100). Relative to the no-vaccination/no-immunity scenario, the corresponding reductions are approximately 14% at the peak, 11% (12%) by year 50 (100). In summary, these results indicate that vaccination provides a sustained additional reduction in malaria burden beyond the model with ITN-use as the only control, with consistent benefits across both endemic and immunologically naive populations. From a public health perspective, these results show that while ITNs constitute an important first level of protection in the community, vaccination constitutes an important the second line of protection.

#### 4.2.3. Global uncertainty and sensitivity analysis

Due to uncertainty, parameter variability, and lack of information on several model parameters, we carry out a global sensitivity analysis on Model (2.1) using Latin Hypercube Sampling (LHS) with a sample size of 1, 000 assuming a 25% variation above and below the baseline parameter values reported in Tables 2. Partial Rank Correlation Coefficients (PRCCs) are calculated to identify the most influential parameters driving malaria transmission and control dynamics systematically. The PRCCs thus obtained give a sense of the relative importance of parameters for malaria transmission quantified through two metrics–peak incidence and the control reproduction number (ℛ_0_). Statistically significant parameters depicted in Figure 5, indicate that the background mosquito mortality rate (*µ*_*v*0_) along with the maximum mosquito biting rate (*β*_max_) are the top two parameters that impact both response metrics, with *µ*_*v*0_ (*β*_max_) negatively (positively) correlated with both metrics. Secondly, with the exception of the waning rate of naturally acquired immunity (*ω*_*r*_) and parasite clearance rate (*ρ*_*h*_)- with the former having a significant negative response on ℛ_0_ but insignificant on the peak incidence, peak incidence and ℛ_0_ are highly sensitive to *α* and *E*_0_ with negative responses on both response metrics. It should be mentioned here that *ρ*_*h*_ is one of the strongly negatively impacting parameters that moves individuals rapidly from the infectious to susceptible state, thus reducing the exposure period to infect mosquitoes. Therefore, from the point of view of public health policy, this parameter emphasizes the importance of accurate and timely diagnosis and treatment in malaria control. On the other hand, the intrinsic mosquito birth rate (*r*_*v*_), the mosquito carrying capacity (*K*_*v*_), the probabilities of transmission from human-to-vector and vector-to-human (*p*_*hv*_ and *p*_*vh*_) and modification factor for infection to the partially immune class (*θ*) are highly impactful to both ℛ_0_ and peak incidence with a positive effect. Other statistically significant parameters associated with reductions in both outcomes, albeit to a lesser extent, include the baseline ITN coverage (*C*_0_), personal protection efficacy (*ε*_*p*_), mosquito-killing efficacy (*ε*_*e*_), and the ITN-induced mosquito mortality rate (*µ*_*v*1_). The shape parameter (*n*) and rate of ITN efficacy waning rate (*σ*_*e*_) are both positively affecting the uncertainty in both measures. Surprisingly, the recovery rates are positively correlated with the control reproduction number and negatively correlated with the peak incidence; in other words, an increase in recovery rates moves individuals out of the symptomatic infectious into the recovered class, where they remain infectious to mosquitoes for a prolonged period because of the slow rate of loss of immunity (*ω*_*r*_). From the perspective of public health policy, these findings show that disease control should target vector biting behaviour and optimizing human protection. Consequently, public health strategies should prioritize scaling up and sustaining ITN coverage with effective nets alongside other targeted vector control to reduce mosquito biting. The significant negative sensitivity of vaccine efficacy highlights the fact that improving on immunization with highly effective vaccines serves as a powerful, complementary intervention within an integrated malaria control framework.

**Figure 5.**
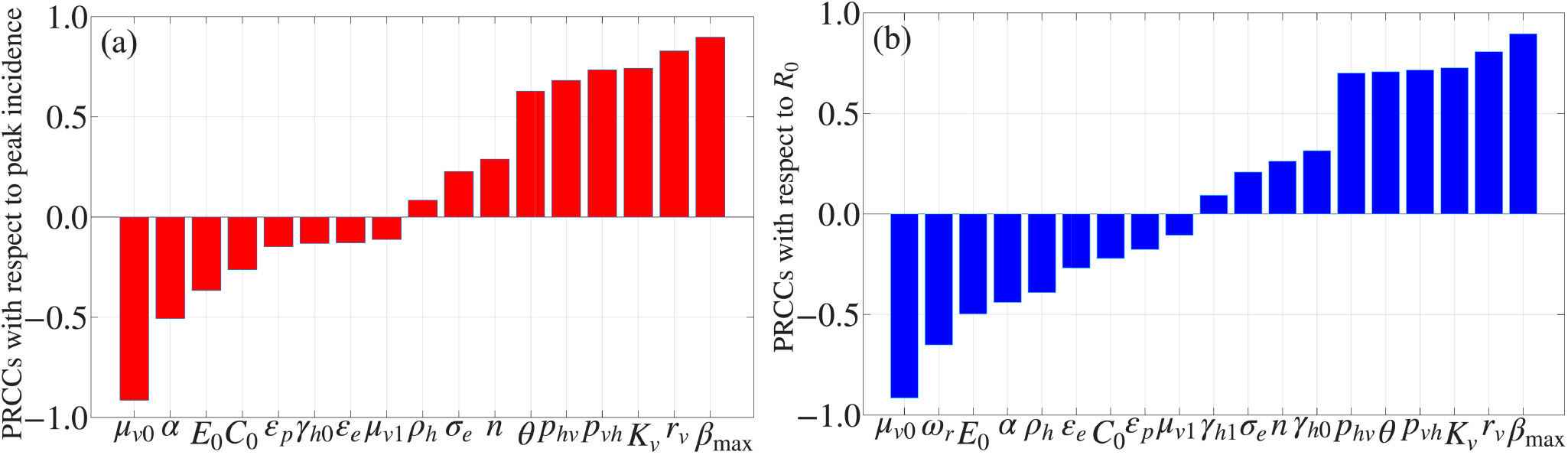
Partial Rank Correlation Coefficients (PRCCs) illustrating the sensitivity of the (a) peak incidence and (b) control reproduction number (ℛ_0_) of Eqs. (2.1) to the parameters. The global sensitivity analysis, carried out through Latin Hypercube Sampling with 1,000 samples (±25% variation), identifies the background mosquito death rate (*µ*_*v*0_) and maximum mosquito biting rate (*β*_max_) as the most significant drivers across both metrics. The other parameter values are given in Table 2.

#### 4.2.4. Identifying single malaria intervention thresholds

To identify threshold levels of various model parameters through which interventions can be assessed, the control reproduction number is plotted as a function of single model parameters under (0.7*β*_max_), baseline (1.0*β*_max_), and high (1.3*β*_max_) mosquito biting rates. Figure 6 depicts the threshold values of these interventions required to reduce the reproduction number below one under different mosquito biting intensities. Under low transmission, modest improvements in many of these control measures are sufficient to interrupt transmission. In particular, the reproduction number can be brought below unity for ITN efficacy greater than 35% (Fig. 6(a)), ITN coverage of about 85% (Fig. 6(b)), ITN intensity of about 9.5 (Fig. 6(c)), vaccination rate of about 0.24 (Fig. 6(d)), vaccine efficacy by about 30% (Fig. 6(e)), and vaccine coverage of about 37% (Fig. 6(f)), whereas the ITN-induced mosquito mortality should be greater than 17 (Fig. 6(g)) and the relative infectiousness of the partially immune class needs to be less than 33% (Fig. 6(h)). That is, the required minimum ITN efficacy for containment changes from about 35% to about 46% (about 57%) under baseline (high) mosquito biting intensity (Fig. 6(a)), while the corresponding required ITN-induced mosquito mortality changed from 17 to about 32 and, to a slightly lesser extent, 37 (Fig. 6(g)). Hence, in low-transmission settings, containing malaria can be achieved through improvements in a range of individual interventions. In contrast, several interventions that are effective in the low-transmission scenario cease to be sufficient as biting intensity increases. In particular, the minimum ITN efficacy required for control rises from 35% to 46% and 57% under baseline and high biting intensities, respectively, (Fig. 6(a)), while the corresponding ITN-induced mosquito mortality threshold increases from 17 to 32 and 37 (Fig. 6(g)). At the same time, the infectiousness modification parameter of partially immune individuals must be below 16% and 10% under baseline and high transmission conditions, respectively (Fig. 6(h)). To summarize, with increasing biting intensities, it becomes increasingly difficult to achieve malaria control. On the other hand, improvement in one control measure alone cannot reduce the reproduction number below one under the baseline and high biting intensity scenarios. Instead, coordinated improvements across multiple interventions are required to reduce ℛ_0_ below one.

**Figure 6.**
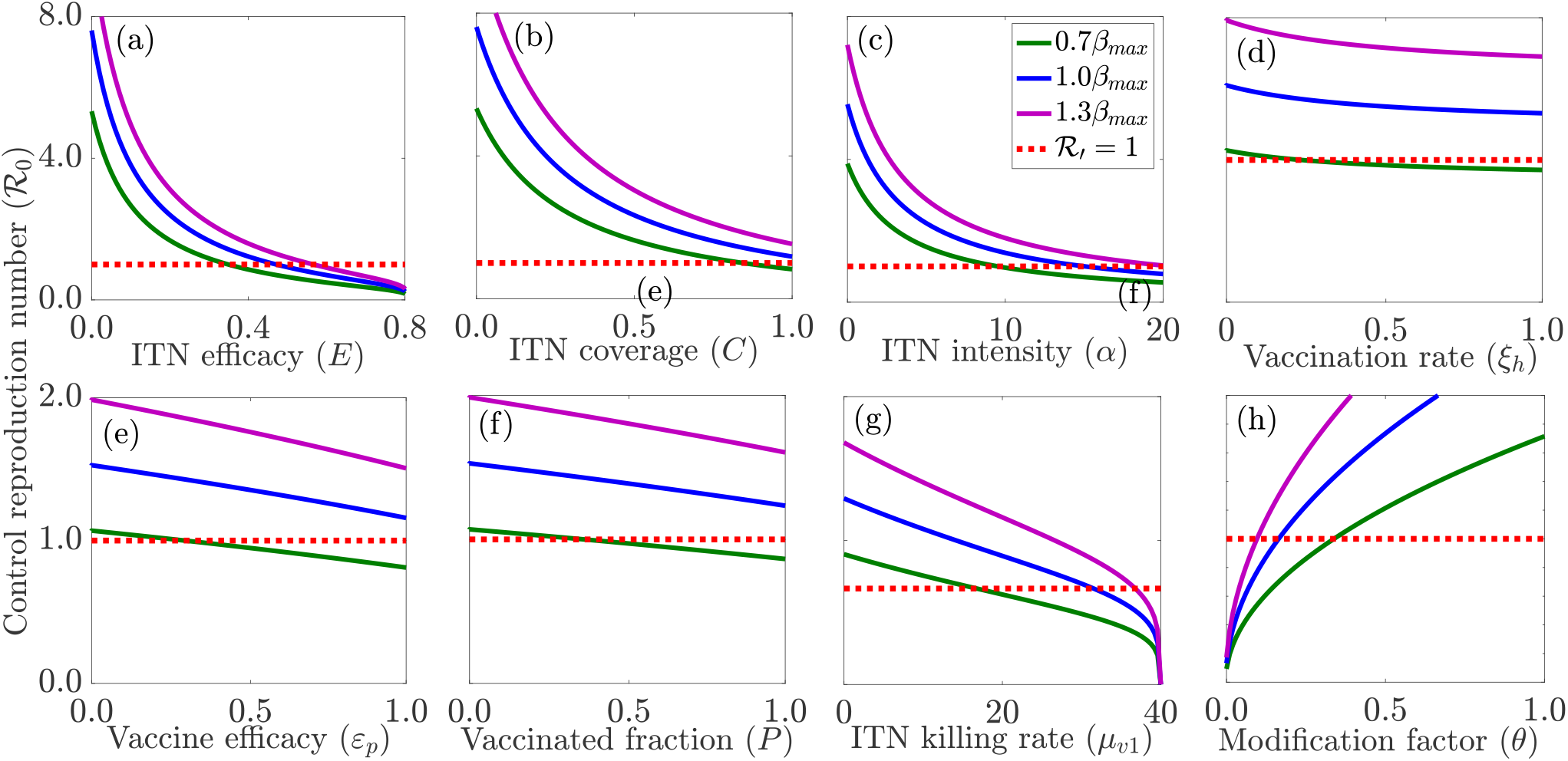
Minimum intervention levels required for malaria control under varying maximum mosquito biting intensities. The subplots depict the dependence of ℛ_0_ on (a) ITN efficacy (*E*), (b) ITN coverage (*C*), (c) ITN intensity (*α*), (d) vaccination rate (*ξ*_*h*_), (e) vaccine efficacy (*ε*_*p*_), (f) vaccination coverage (*P*), (g) ITN-induced mosquito mortality rate (*µ*_*v*1_), and (h) the infectiousness modification factor (*θ*) of partially immune humans for low (0.7*β*_max_), baseline (1.0*β*_max_), and high (1.3*β*_max_) mosquito biting rates. Dotted lines indicate the threshold ℛ_0_ = 1. Values of the other parameters used are given in Tables 2.

#### 4.2.5. Assessing the combined impacts of ITN and vaccination on the reproduction number

To assess the synergistic impact of interventions on malaria control, we carry out a sensitivity analysis of the control reproduction number (ℛ_0_), as a function of pairs of parameters through which interventions can be implemented. By mapping ℛ_0_ in coupled two-dimensional parameter spaces, we obtain the threshold boundaries where ℛ_0_ = 1 and the best combinations of ITN and vaccine coverage to control malaria. The contour plots in Fig. 7 reveal a strong synergistic effect between ITN use and vaccination. Using the baseline parameter values we cannot contain malaria, even with complete ITN use and full vaccination coverage suggesting that combined intervention effectiveness is insufficient to bring transmission below the epidemic threshold (Fig. 7(a)). Any improvement in either intervention shifts the control boundary substantially. For instance, with ITN efficacy increased to 75% from its baseline value (Fig. 7(b)), malaria can be controlled if at least 36% of the population is vaccinated, given complete ITN use, or at least 89% of the population uses ITNs, given complete vaccination. Similarly, with vaccine efficacy increased to 75% from its baseline value (Fig. 7(c)), the minimum vaccine coverage needed for control under full ITN use decreases to 53%, and under full vaccination, the minimum ITN coverage needed for control decreases to 66%. In this case, full ITN use needs a minimum vaccination coverage of 17%, and full vaccination only needs a minimum ITN coverage of 57%. Therefore, controlling malaria depends on both coverage and efficacy of interventions such as ITN use and vaccination. Increasing ITN or vaccine efficacy decreases the coverage needed to interrupt transmission, with the most significant decreases in coverage needed when both are improved.

**Figure 7.**
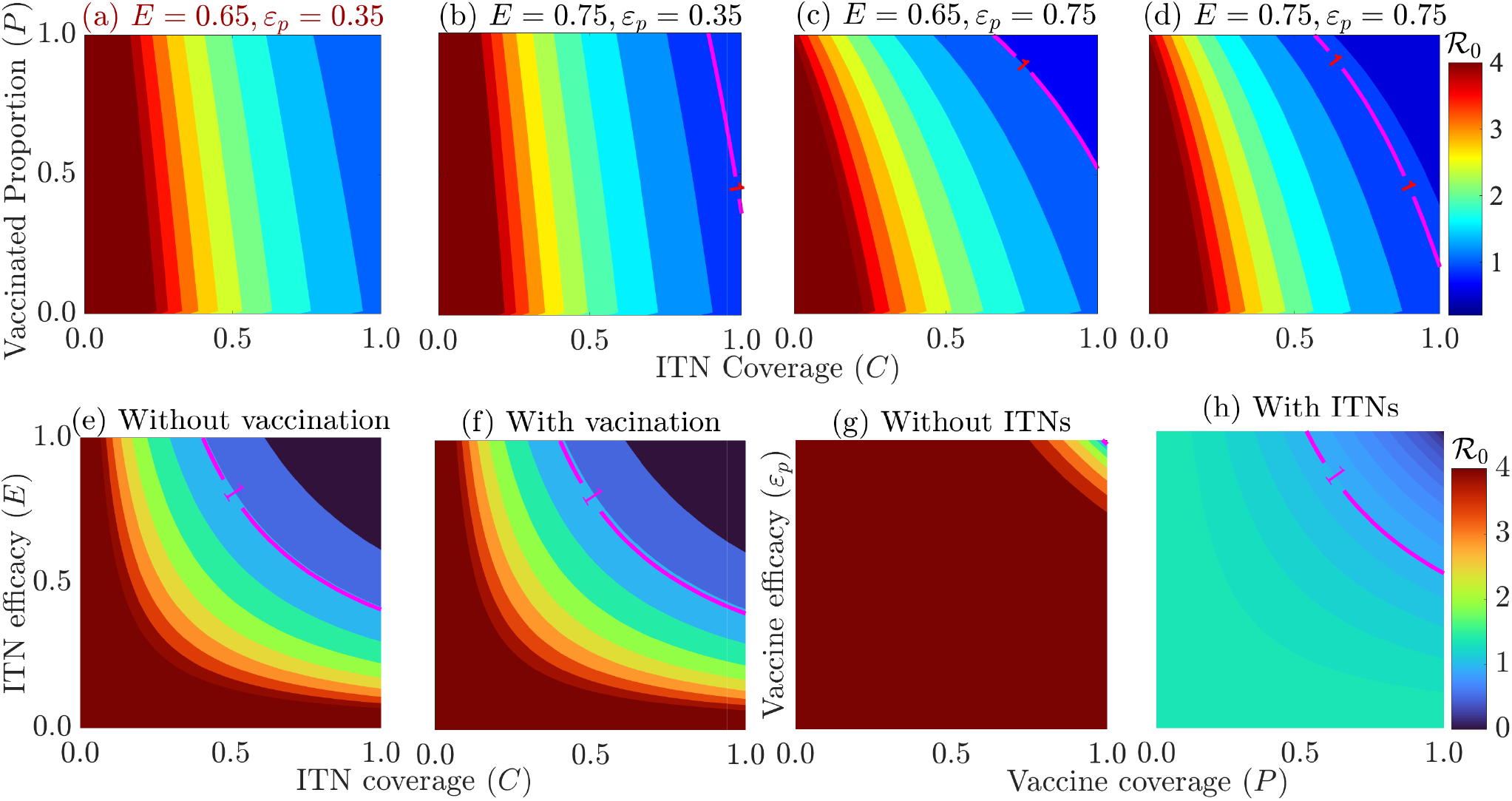
Containing malaria depends on both the coverage and efficacy of ITNs and vaccination. Under baseline conditions ((a)), transmission cannot be interrupted even with universal ITN use and vaccination. Increasing ITN efficacy ((b)) or vaccine efficacy ((c)) shifts the containment threshold, making elimination feasible at attainable coverage levels. The greatest reduction in transmission occurs when both interventions are highly effective ((d)), substantially lowering the coverage required for containment. (e)-(f) Minimum ITN efficacy required to reduce ℛ_0_ < 1 under universal community usage, with ((e)) or without ((f)) vaccination. (g) Required minimum vaccine efficacy under universal vaccination in the absence of ITN use. (h) Required minimum vaccine efficacy under universal vaccination when paired with universal ITN use. These results demonstrate the synergistic benefits of combining vector control and vaccination and highlight intervention efficacy as a key determinant of malaria elimination feasibility. The other parameter values are listed in Tables 2.

While these optimum coverages present an ideal scenario, the absolute minimum efficacies of each intervention, in the absence and presence of the other, provide absolute limits to transmission control. Reducing the reproduction number below one requires an approximate minimum ITN efficacy of 40%, assuming universal community usage, regardless of vaccination status (Fig. 7(e) and (f)). In the absence of ITNs, relying solely on universal vaccination requires a minimum vaccine efficacy of ≈ 99% to interrupt transmission (Fig. 7(g)). But with ITN use accounted for, the required minimum vaccine efficacy for a fully vaccinated population drops to about 53% (Fig. 7(h)). This provides context for evaluating current vaccine candidates. For example, when combined with ITNs, a moderately effective vaccine such as RTS,S/AS01 (*ε*_*p*_ ∈ [0.30, 0.40]) will not allow disease control, even if the entire population is targeted for vaccination. On the other hand, combining ITNs with the more effective R21/Matrix-M vaccine (*ε*_*p*_ ∈ [0.66, 0.75]) allows disease control at coverage levels between 70% and 81% of the target population. These results strongly favor integrated malaria control strategies, which combine high-effective vector control with a vaccination campaign, over single-measure control strategies.

## 5. Discussion and conclusion

Mathematical modeling has provided useful insights into malaria transmission and the impact of control measures, particularly ITNs. However, by reducing the risk of infection, ITN-use reduces the level of immunity that the population can acquire. Here, we model the interplay between ITN-use, vaccination, and immunity dynamics with a framework in which ITN coverage and efficacy as time-varying states and link immunity acquisition according to infection exposure and vaccination. This framework allows us to assess whether ITN-use is beneficial or detrimental to malaria control and to identify optimum ITN and vaccination coverage levels that are appropriate for containing malaria. The results obtained offer insights into the balance between reducing disease burden and maintaining immunity in malaria-endemic settings.

Simulations of the model confirm the fact that under baseline conditions, ℛ_0_ is as a classical threshold that separates disease elimination (when ℛ_0_ < 1) from persistence (when ℛ_0_ > 1). However, for high malaria mortality levels, a backward bifurcation becomes possible. This agrees with the malaria modeling literature discussed in [15, 16, 22, 23], showing that nonlinear processes could render ℛ_0_ a little helpful as a sole epidemiological explanatory variable. Interpreted epidemiologically, containing malaria cannot be guaranteed by assuring the condition ℛ_0_ < 1. Instead, the initially present infection level is important, so that further efforts at reducing transmission levels below some lower critical threshold is required. In addition, the results suggest that increasing ITN coverage reduces malaria incidence and population-level immunity, especially at higher coverages. This is because the reduced contact between humans and mosquitoes, increased mortality of mosquitoes, and reduced mosquito population reduce transmission. Therefore, the benefits of protection exceed those of acquired immunity. From a public health perspective, increasing ITN coverage is important for reducing malaria burden and combining ITN-use with vaccination will lead to even more reduction in malaria burden and improved population-level immunity.

Analytical establishment of the global asymptotic stability of the extinction, mosquito-free, and disease-free equilibria provides a rigorous characterization of the threshold dynamics of the model within the epidemiologically relevant parameter regime. Although numerical bifurcation analysis reveals the theoretical possibility of a backward bifurcation, in our model the phenomenon arises only for unrealistically large values of the disease-induced mortality rate; values that lie far outside the biologically plausible range for malaria. Consequently, it is not certain whether this phenomenon will still exist under realistic epidemiological conditions. Therefore, the global stability results we established in Theorem 3.3 remain the appropriate characterization of the system’s dynamics in practice. Nevertheless, the emergence of a backward bifurcation under extreme parameter values highlights the rich nonlinear structure of our model and suggests that a complete analytical characterization of all possible bifurcation regimes warrants further investigation.

A global sensitivity analysis of the model indicates that the vector-human contact parameter sets and intervention effectiveness parameters are the drivers of malaria dynamics. Specifically, uncertainty and variability in entomological parameters such as the mosquito biting rate, mosquito population growth rate, and carrying capacity will trigger the greatest positive uncertainty and variability on malaria peak incidence and ℛ_0_, while uncertainty and variability in mosquito mortality and intervention-related parameters such as ITN protection coverage and efficacy will lead to the greatest negative uncertainty and variability on these metrics. Additionally, there is higher uncertainty in predicting the level of contact with mosquitoes and their general ecology than in measures of control related to protection against bites. Hence, the possible optimum control strategy is contact reduction through ITN-use, mosquito population reduction, and host protection by vaccination.

A comprehensive local sensitivity analysis of the model reveals that malaria control is highly sensitive to the maximum biting rate of mosquitoes. In particular, control under low transmission conditions is achievable with ITNs, vaccination, or vector controls at baseline level. However, at higher maximum biting rates, even with further intensified control measures, such single measures are not sustainable given the intensification of biting rate. On the other hand, ITN and vaccine combinations prove to be highly synergistic because any increase in the efficacy of one of these interventions reduces the coverage threshold of the other intervention. Specifically, for high efficacy combinations, control is possible at realistic coverage, while for low-efficacy vaccines, control is still not feasible, even at perfect ITN coverage. Thus, under realistic transmission settings, malaria control requires combined interventions involving effective ITNs and highly efficacious vaccination, as neither intervention alone is sufficient in regions with very high biting rates. This is in agreement with the nonlinearity of malaria control and multi-measures plan, as documented recently in modeling studies of malaria dynamics [27, 31].

While this study provides useful insights into the interplay between malaria transmission dynamics, immunity, and ITN-use, several opportunities exist for future refinement. The deterministic differential equation model does not address stochastic variability that may be associated with parameters of the model (e.g., biting and death rates) or random fluctuations in transmission, particularly in low-transmission settings. Also, it does not examine the effects of temperature and precipitation on the dynamics of mosquito populations. The model assumes homogeneous mixing in the population. Relaxing this assumption to account for spatial, demographic, and behavioral heterogeneity may provide a more detailed representation of exposure patterns, immunity acquisition, and ITN-use. Additionally, future studies could account for operational and logistical aspects of ITN distribution, replacement, access, and adherence. Furthermore, accounting for challenges related vaccine allocation, equitable access, uptake, and public trust, can have a significant impact on the effectiveness of vaccination programs and their interaction with acquired immunity. Incorporating these, and other aspects such as compartmentalizing the mosquito population to differentiate questing mosquitoes from resting ones, are avenues of enquiry that are presented elsewhere.

To conclude, this study highlights the fact that malaria control is nonlinear, with ℛ_0_ slightly less than one not sufficient for elimination due to backward bifurcation and persistence driven by residual infection levels. Although ITN use reduces natural immunity by limiting mosquito exposure, this loss is more than offset by the substantial public health gains from reduced mosquito–human contact, increased vector mortality, and reduced transmission. These benefits are strengthened when ITNs are complemented with vaccination, which increases host protection and lowers the intervention thresholds needed for control. Thus, sustained and integrated use of ITNs and vaccines provides the most reliable pathway for durable malaria reduction across transmission settings.

## Data Availability

All data produced in the present study are available upon reasonable request to the authors

## Acknowledgments

AJOT acknowledges the University of the Witwatersrand for support during his postdoctoral fellowship, where this work was initiated. GAN and BMG acknowledge the support from the Cameroon Ministry of Higher Education through the initiative for the modernisation of research in Cameroon’s Higher Education for 2025 and 2026. CNN was supported by the U.S. National Science Foundation under Grant No. DMS-2151870. The authors thank the organizers of the Mathematics in Industry Study Group (MISG) 2025, especially Prof. David Mason and Prof. Neville Fowkes for selecting this topic and problem and for their support throughout the program. The authors gratefully acknowledge the contributions of members of the mosquito dynamics study group, whose insights and engagement helped shape the early development of this research. Preliminary work on this problem began at the workshop through collaborative discussions and problem-solving sessions.

