## Supplementary Information for "The Immunity Paradox of Bed Nets: Why Reducing Exposure Can Still Strengthen Malaria Control"

### Supplementary Information (SI)

### S1. Analysis of the model (2.1) with general functions $f_h(N_h)$ and $f_v(N_v)$

#### *S1.1. Positively invariant and attracting set (Proof of Theorem 3.1)*

Let the biologically feasible domain for the system be $\Omega \subset \mathbb{R}^9_+$. To establish the positive invariance and global attractiveness of $\Omega$ under the general demographic functions $f_h(N_h)$ and $f_v(N_v)$, we analyze the total host population $N_h = S_h+V_h+I_h+R_h$ and total vector population $N_v = S_v+I_v$. Specifically, the eqations for the total human and vector populations are given by $\dot{N}_h = f_h(N_h) - \mu_h N_h - \delta_h(M)I_h \le f_h(N_h) - \mu_h N_h$ and $\dot{N}_v = f_v(N_v) - \mu_v N_v$. If we define the human and mosquito carrying capacities as $N_{h,\max} = \sup\{N_h \ge 0 : f_h(N_h) - \mu_h N_h \ge 0\}$ and $N_{v,\max} = \sup\{N_v \ge 0 : f_v(N_v) - \mu_v N_v \ge 0\}$, then by comparison arguments, $\limsup_{t\to\infty} N_h(t) \le N_{h,\max}$ and $\limsup_{t\to\infty} N_v(t) \le N_{v,\max}$, respectively. Hence, $0 \le S_h, V_h, I_h, R_h \le N_{h,\max}$, $0 \le S_v, I_v \le N_{v,\max}$. The population-level immunity ($M$), ITN-efficacy ($E$), and coverage ($C$), are sequentially bounded via their uniform limits. In particular, noting that $\Psi \le \tau_d$, $V_h \le N_{h,\max}$, and $\Phi \ge \mu_h$, the population-level immunity equation reduces to $\dot{M} \le \tau_d + \tau_p\varepsilon_p N_{h,\max} - \mu_h M$. This implies $\limsup_{t\to\infty} M(t) \le \frac{\tau_d+\tau_p\varepsilon_p N_{h,\max}}{\mu_h} \equiv M_{\max}$. Furthermore, the linear ITN-efficacy equation $\dot{E} = \varepsilon_e E_0 - (\varepsilon_e + \sigma_e)E$ implies $\limsup_{t\to\infty} E(t) = \frac{\varepsilon_e E_0}{\varepsilon_e+\sigma_e} \equiv E_{\max}$. For the ITN-coverage variable ($C$): Since $g(E, N_v)$ is increasing monotonically in $E$ and $N_v$, let $g_{\max} = g(E_{\max}, N_{v,\max})$. The inequality $\dot{C} \le g_{\max}C_0 - (g_{\max} + \sigma_c)C$ yields $\limsup_{t\to\infty} C(t) \le \frac{g_{\max}C_0}{g_{\max}+\sigma_c} \le C_0 \equiv C_{\max}$. The bounded region $\Omega$ is defined as: $\Omega = \{(S_h, V_h, I_h, R_h, M, E, C, S_v, I_v) \in \mathbb{R}^9_+ : 0 \le N_h \le N_{h,\max}, 0 \le E \le E_{\max}, 0 \le C \le C_{\max}, 0 \le M \le M_{\max},\ 0 \le N_v \le N_{v,\max}\}$.

**Theorem S1.1.** *The region $\Omega$ is positively invariant and globally attracting for the model.*

*Proof.* Evaluating the vector field on the boundary faces of $\Omega$ shows that all normal derivatives point inward or are tangent to $\partial\Omega$ for all $t \ge 0$. Since every state variable is asymptotically bounded by its respective supremum via comparison, $\Omega$ is a global absorbing set in $\mathbb{R}^9_+$. □

#### *S1.2. Existence of disease-free equilibria*

Equilibria of the full model are obtained by setting the derivatives on the left hand sides to zero and solving the resulting system of algebraic equations. At disease-free equilibria, $I_h = I_v = 0$, which forces $\lambda_{vh} = \lambda_{hv} = 0$, and $R = 0$. Substituting these in the system and solving the resulting system leads to up to three possible equilibria to the model in terms of the general demographic functions $f_h(N_h)$ and $f_v(N_v)$.

*1. The trivial equilibrium ($\mathcal{E}_0$).* This equilibrium corresponds to the total extinction of both human and vector populations. It is given by

$$\mathcal{E}_0 = (S_h^*, V_h^*, I_h^*, R_h^*, M^*, E^*, C^*, S_v^*, I_v^*) = \left(0, 0, 0, 0, 0, \frac{\varepsilon_e E_0}{\varepsilon_e + \sigma_e}, 0, 0, 0\right).$$

This equilibrium always exists for any choice of $f_h$ and $f_v$ provided that $f_h(0) = 0$ and $f_v(0) = 0$.

*2. The mosquito-free equilibrium ($\mathcal{E}_m$).* This equilibrium represents a state where the human population persists and reaches a demographic steady state, but the vector population goes extinct. It is given by

$$\mathcal{E}_m = (S_h^*, V_h^*, 0, 0, M^*, E^*, 0, 0, 0),$$

where the non-zero component expressions are given explicitly by:

$$S_h^* = \frac{\omega_p + \mu_h}{\xi_h + \omega_p + \mu_h} N_h^*, \quad V_h^* = \frac{\xi_h}{\xi_h + \omega_p + \mu_h} N_h^*, \quad M^* = \frac{\tau_p \varepsilon_p \xi_h N_h^*}{\mu_h(\xi_h + \omega_p + \mu_h) + \omega_p \xi_h} \quad E^* = \frac{\varepsilon_e E_0}{\varepsilon_e + \sigma_e},$$

and $N_h^*$ is the positive solution of the equation: $f_h(N_h^*) = \mu_h N_h^*$. A biologically meaningful state requires that a unique, positive real solution ($N_h^* > 0$) exists for the equation above.

*3. The full disease-free equilibrium ($\mathcal{E}_{DFE}$).* This represents the equilibrium where both human host and vector populations coexist in the absence of the disease. It is given by

$$\mathcal{E}_{DFE} = (S_h^*, V_h^*, 0, 0, M^*, E^*, C^*, S_v^*, 0),$$

where the non-zero state expressions are:

$$\begin{aligned} S_h^* &= \frac{\omega_p + \mu_h}{\xi_h + \omega_p + \mu_h} N_h^*, \quad V_h^* = \frac{\xi_h}{\xi_h + \omega_p + \mu_h} N_h^*, \quad M^* = \frac{\tau_p \varepsilon_p \xi_h N_h^*}{\mu_h(\xi_h + \omega_p + \mu_h) + \omega_p \xi_h}, \\ E^* &= \frac{\varepsilon_e E_0}{\varepsilon_e + \sigma_e}, \quad C^* = \frac{\varphi E^* N_v^* C_0}{\varphi E^* N_v^* + \sigma_c(1 + \phi E^* N_v^*)}, \quad S_v^* = N_v^*, \end{aligned}$$

where $N_h^*$ is the positive solution to the host demographic equation: $f_h(N_h^*) = \mu_h N_h^*$ and $N_v^*$ is the positive solution of the equation: $f_v(N_v^*) = \mu_v N_v^*$. For $\mathcal{E}_{DFE}$ to exist in the positive orthant, the growth functions must allow persistence from low densities. That is, $f_h'(0) > \mu_h$ and $f_v'(0) > \mu_v$ (allowing vector existence). *We can also use $\mathcal{T}_h > 1, \mathcal{T}_h = f_h'(0)/\mu_h$ and $\mathcal{T}_v > 1, \mathcal{T}_v = f_v'(0)/\mu_v$.*

#### *S1.3. Computation of the control and basic reproduction numbers of the full model*

Let us assume $\mathcal{T}_h$ and $\mathcal{T}_v$ greater than one and define by $\mathcal{F}$ and $\mathcal{V}$ the vectors of the new infections and the transfer terms of the model, respectively.

$$\mathcal{F} = \begin{pmatrix} \lambda_{vh}(S_h + (1 - \varepsilon_p)V_h) \\ 0 \\ \lambda_{hv} S_v \end{pmatrix} \text{ and } \mathcal{V} = \begin{pmatrix} (\gamma_h(M) + \delta_h(M) + \rho_h + \mu_h) I_h \\ -\gamma_h I_h + (\omega_r + \mu_h) R_h \\ \mu_v I_v \end{pmatrix}$$

The Jacobian matrix $F$ and $V$ of $\mathcal{F}$ and $\mathcal{V}$ at the DFE, $\mathcal{E}_1$, are given by

$$F = \begin{pmatrix} 0 & 0 & \dfrac{\beta^* p_{vh}[S_h^* + (1 - \varepsilon_p)V_h^*]}{N_h^*} \\ 0 & 0 & 0 \\ \dfrac{\beta^* p_{hv} S_v^*}{N_h^*} & \dfrac{\beta^* \theta p_{hv} S_v^*}{N_h^*} & 0 \end{pmatrix} \text{ and}$$

$$V = \begin{pmatrix} [\gamma_h(M^*) + \delta_h(M^*) + \rho_h + \mu_h] & 0 & 0 \\ -\gamma_h & (\omega_r + \mu_h) & 0 \\ 0 & 0 & \mu_v^* \end{pmatrix}, \text{ respectively.}$$

From the $V$-matrix, we obtain

$$V^{-1} = \begin{pmatrix} \dfrac{1}{[\gamma_h(M^*) + \delta_h(M^*) + \rho_h + \mu_h]} & 0 & 0 \\ \dfrac{\gamma_h}{[\gamma_h(M^*) + \delta_h(M^*) + \rho_h + \mu_h](\omega_r + \mu_h)} & \dfrac{1}{\omega_r + \mu_h} & 0 \\ 0 & 0 & \dfrac{1}{\mu_v^*} \end{pmatrix} \text{ and the next generation matrix}$$

$$FV^{-1} = \begin{pmatrix} 0 & 0 & \dfrac{\beta^* p_{vh}[S_h^* + (1-\varepsilon_p)V_h^*]}{N_h^* \mu_v^*} \\ 0 & 0 & 0 \\ \dfrac{\beta^* p_{hv} S_v^* (\omega_r + \mu_h + \theta\gamma_h)}{N_h^*[\gamma_h(M^*) + \delta_h(M^*) + \rho_h + \mu_h](\omega_r + \mu_h)} & \dfrac{\beta^* \theta p_{hv} S_v^*}{N_h^*(\omega_r + \mu_h)} & 0 \end{pmatrix}$$

The reproduction number (i.e., the spectral radius of the matrix, $FV^{-1}$) is

$$\mathcal{R}_0 = \sqrt{\underbrace{\frac{\beta(E^*,C^*)p_{vh}}{\gamma_h(M^*) + \delta_h(M^*) + \rho_h + \mu_h}\left[\frac{S_h^* + (1-\varepsilon_p)V_h^*}{N_h^*}\right]}_{\text{mosquito}\to\text{human}} \cdot \underbrace{\frac{\beta(E^*,C^*)p_{hv}\, N_v^*}{N_h^*\, \mu_v(E^*,C^*)}\left(1 + \frac{\theta\gamma_h(M^*)}{\omega_r + \mu_h}\right)}_{\text{human}\to\text{mosquito}}}, \quad \text{(S1.1)}$$

where $\mathcal{R}_0 = R_0(M^*)\sqrt{1-\varepsilon_p P^*}$, $R_0(M^*) = \sqrt{\frac{\beta(E^*,C^*)p_{vh}}{[\gamma_h(M^*)+\delta_h(M^*)+\rho_h+\mu_h]} \cdot \frac{\beta(E^*,C^*)p_{hv}\, N_v^*}{N_h^*\, \mu_v(E^*,C^*)}\left(1 + \frac{\theta\gamma_h(M^*)}{\omega_r+\mu_h}\right)}$, with $\beta(E^*,C^*) = \frac{\beta_{max}}{1+\alpha E^* C^*}$, $\mu_v^* := \mu_v(E^*,C^*) = \mu_{v0} + \mu_{v1}E^*C^*$, $\gamma_h(M^*) = \gamma_{h0} + \frac{\gamma_{h1}M^*}{1+M^*}$, $\delta_h(M^*) = \delta_{h0} + \frac{\delta_{h1}}{1+M^*}$. $P^* = V_h^*/N_h^*$ is the vaccine coverage. Since $N_h^* = S_h^* + V_h^*$, $[S_h^* + (1-\varepsilon_p)V_h^*]/N_h^* = 1 - \varepsilon_p P^*$.

#### *S1.4. Local stability of disease-free equilibria*

To study the local stability of the three disease-free equilibria ($\mathcal{E}_0, \mathcal{E}_m$, and $\mathcal{E}_{DFE}$), we linearize the full model system at each of the equilibria. In particular, if $X = (S_h, V_h, I_h, R_h, M, E, C, S_v, I_v)^T \in \mathbb{R}_+^9$. The nonlinear system is governed by $\dot{X} = F(X)$. At any disease-free state, the infected classes vanish (i.e., $I_h = R_h = I_v = 0$), which implies $\lambda_{vh} = \lambda_{hv} = 0$, $\Psi = 0$, and $g(E, N_v) = g(E, S_v)$. Evaluating the partial derivatives $A_{ij} = \frac{\partial F_i}{\partial X_j}$ under these conditions yields the general disease-free Jacobian structure:

$$\mathcal{J} = \begin{pmatrix} A_{11} & A_{12} & A_{13} & A_{14} & 0 & 0 & 0 & 0 & A_{19} \\ A_{21} & A_{22} & 0 & 0 & 0 & 0 & 0 & 0 & A_{29} \\ 0 & 0 & A_{33} & 0 & 0 & 0 & 0 & 0 & A_{39} \\ 0 & 0 & A_{43} & A_{44} & 0 & 0 & 0 & 0 & 0 \\ 0 & A_{52} & A_{53} & A_{54} & A_{55} & 0 & 0 & 0 & A_{59} \\ 0 & 0 & 0 & 0 & 0 & A_{66} & 0 & 0 & 0 \\ 0 & 0 & 0 & 0 & 0 & A_{76} & A_{77} & A_{78} & 0 \\ 0 & 0 & A_{83} & A_{84} & 0 & 0 & 0 & A_{88} & 0 \\ 0 & 0 & A_{93} & A_{94} & 0 & 0 & 0 & 0 & A_{99} \end{pmatrix},$$

where the non-zero general entries are given by

$$
\begin{aligned}
&A_{11} = f_h'(N_h) - (\xi_h + \mu_h), && A_{12} = f_h'(N_h) + \omega_p, && A_{13} = f_h'(N_h) + \rho_h,\\
&A_{14} = f_h'(N_h) + \omega_r, && A_{19} = -\tfrac{\beta(E,C)p_{vh}S_h}{N_h}, && A_{21} = \xi_h,\\
&A_{22} = -(\omega_p + \mu_h), && A_{29} = -(1-\varepsilon_p)\tfrac{\beta(E,C)p_{vh}V_h}{N_h}, && A_{33} = -(\gamma_h(M) + \delta_h(M) + \rho_h + \mu_h),\\
&A_{39} = \tfrac{\beta(E,C)p_{vh}[S_h+(1-\varepsilon_p)V_h]}{N_h}, && A_{43} = \gamma_h(M), && A_{44} = -(\omega_r + \mu_h),\\
&A_{52} = \tau_p\varepsilon_p - \omega_p\tfrac{M(N_h-V_h)}{N_h^2}, && A_{53} = -\delta_h(M)M, && A_{54} = -\omega_r\tfrac{M}{N_h},\\
&A_{55} = -\left(\mu_h + \omega_p\tfrac{V_h}{N_h}\right), && A_{59} = \tfrac{\tau_d\beta(E,C)p_{vh}(S_h+(1-\varepsilon_p)V_h)}{dN_h}, && A_{66} = -(\varepsilon_e + \sigma_e),\\
&A_{76} = \tfrac{\partial g}{\partial E}(C_0 - C), && A_{77} = -g(E,S_v) - \sigma_c, && A_{78} = \tfrac{\partial g}{\partial S_v}(C_0 - C),\\
&A_{83} = -\tfrac{\beta(E,C)p_{hv}S_v}{N_h}, && A_{84} = -\tfrac{\beta(E,C)p_{hv}\theta S_v}{N_h}, && A_{88} = f_v'(S_v) - \mu_v,\\
&A_{93} = \tfrac{\beta(E,C)p_{hv}S_v}{N_h}, && A_{94} = \tfrac{\beta(E,C)p_{hv}\theta S_v}{N_h}, && A_{99} = -\mu_v.
\end{aligned}
$$

The rows/columns 5, 6, 7, and 8 are decoupled from the remaining system. Hence, we have the direct eigenvalues $\lambda_1 = A_{55} = -\left(\mu_h + \omega_p\frac{V_h}{N_h}\right) < 0, \lambda_2 = A_{66} = -(\varepsilon_e + \sigma_e) < 0, \lambda_3 = A_{77} = -g(E,S_v) - \sigma_c < 0, \lambda_4 = A_{88} = f_v'(S_v) - \mu_v < 0$, if $f_v'(S_v^*) < \mu_v$. Since all but one of these direct eigenvalues are negative, local stability of the equilibria ($\mathcal{E}_0$, $\mathcal{E}_m$, and $\mathcal{E}_{DFE}$) can be established using $\lambda_4 = f_v'(S_v) - \mu_v$, and the following reduced $5\times 5$ matrix that is obtained by eliminating the rows and columns of $\mathcal{J}$ that correspond to the direct eigenvalues:

$$
\mathcal{J}_{\text{red}} = \begin{pmatrix} A_{11} & A_{12} & A_{13} & A_{14} & A_{19}\\ A_{21} & A_{22} & 0 & 0 & A_{29}\\ 0 & 0 & A_{33} & 0 & A_{39}\\ 0 & 0 & A_{43} & A_{44} & 0\\ 0 & 0 & A_{93} & A_{94} & A_{99} \end{pmatrix}.
$$

*S1.4.1. Local stability of the trivial equilibrium $\mathcal{E}_0$*

To study the local stability of the extinction equilibrium ($\mathcal{E}_0 = (0,0,0,0,0,E^*,0,0,0)$), we evaluate the coefficients $A_{ij}$ at $\mathcal{E}_0$. In addition to the eigenvalues above, this further isolates $\lambda_5 = A_{99} = -\mu_{v0} < 0, \lambda_6 = A_{33} = -(\gamma_{h0} + \delta_{h0} + \rho_h + \mu_h) < 0$, and $\lambda_7 = A_{44} = -(\omega_r + \mu_h) < 0$. The remaining two eigenvalues can be determined from the further reduced $2\times 2$ matrix:

$$
\mathcal{J}_{\text{dem}} = \begin{pmatrix} f_h'(0) - (\xi_h + \mu_h) & f_h'(0) + \omega_p\\ \xi_h & -(\omega_p + \mu_h) \end{pmatrix}.
$$

For local stability of $\mathcal{E}_0$, we require $\lambda_4 = f_v'(0) - \mu_{v0} < 0$, $\text{Tr}(\mathcal{J}_{\text{dem}}) < 0$, and $\text{Det}(\mathcal{J}_{\text{dem}}) > 0$. But $\text{Tr}(\mathcal{J}_{\text{dem}}) = (f_h'(0) - \mu_h) - (\xi_h + \omega_p + \mu_h)$ and $\text{Det}(\mathcal{J}_{\text{dem}}) = [f_h'(0) - (\xi_h + \mu_h)][-(\omega_p + \mu_h)] - \xi_h[f_h'(0) + \omega_p] = (\xi_h + \omega_p + \mu_h)(\mu_h - f_h'(0)) \implies \text{Tr}(\mathcal{J}_{\text{dem}}) < 0$ and $\text{Det}(\mathcal{J}_{\text{dem}}) > 0$ if $f_h'(0) < \mu_h$. Thus, if $f_h'(0) < \mu_h$ and $f_v'(0) < \mu_{v0}$, all eigenvalues of $\mathcal{J}$ evaluated at $\mathcal{E}_0$ are negative or have negative real parts (if they are complex). Therefore, the trivial equilibrium $\mathcal{E}_0 = (0,0,0,0,0,E^*,0,0,0)$ is locally asymptotically stable (LAS) if $f_h'(0) < \mu_h$ and $f_v'(0) < \mu_{v0}$.

*S1.4.2. Local stability of the mosquito-free equilibrium ($\mathcal{E}_m$)*

At the mosquito-free equilibrium ($\mathcal{E}_m$), $N_h^* = S_h^* + V_h^* > 0$, and $S_v^* = N_v^* = 0$. Evaluating the Jacobian ($\mathcal{J}$) or the reduced Jacobian ($\mathcal{J}_{\text{red}}$) at $\mathcal{E}_m$, yields $A_{78} = A_{83} = A_{84} = A_{93} = A_{94} = 0$, the same eigenvalues $\lambda_j, j = 1,2,3,\ldots,7$ in Section S1.4.1, and the reduced $2\times 2$ matrix

$$
\mathcal{J}_{\text{human}} = \begin{pmatrix} f_h'(N_h^*) - (\xi_h + \mu_h) & f_h'(N_h^*) + \omega_p\\ \xi_h & -(\omega_p + \mu_h) \end{pmatrix}.
$$

Applying the Trace-Determinant criterion, we have: $\text{Tr}(\mathcal{J}_{\text{dem}}) = [f_h'(N_h^*) - \mu_h] - (\xi_h + \omega_p + \mu_h)$ and $\text{Det}(\mathcal{J}_{\text{dem}}) = (\xi_h + \omega_p + \mu_h)(\mu_h - f_h'(N_h^*)) \implies \text{Tr}(\mathcal{J}_{\text{dem}}) < 0$ and $\text{Det}(\mathcal{J}_{\text{dem}}) > 0$ if $f_h'(N_h^*) < \mu_h$. But, this is a standard and mathematically true condition since the human population is at its ecological carrying capacity. Hence, if

$\lambda_4 = f_v'(0) - \mu_{v0} < 0$, all eigenvalues of $\mathcal{J}$ evaluated at $\mathcal{E}_m$ are negative or have negative real parts (if they are complex). Therefore, $\mathcal{E}_m$ is locally asymptotically stable if $f_v'(0) < \mu_{v0}$.

*S1.4.3. Local stability of the full disease-free equilibrium ($\mathcal{E}_{DFE}$)*

The reduced matrix ($\mathcal{J}_{\text{red}}$) evaluated at the full disease-free equilibrium ( $\mathcal{E}_{DFE}$) can be expressed as the upper triangular block matrix:

$$\mathcal{J}_{\text{red}} = \begin{pmatrix} \mathcal{A} & \mathcal{B} \\ 0 & \mathcal{C} \end{pmatrix}, \quad \text{where } \mathcal{A} = \begin{pmatrix} A_{11} & A_{12} \\ A_{21} & A_{22} \end{pmatrix}, \quad \mathcal{B} = \begin{pmatrix} A_{13} & A_{14} & A_{19} \\ 0 & 0 & A_{29} \end{pmatrix} \text{ and } \mathcal{C} = \begin{pmatrix} A_{33} & 0 & A_{39} \\ A_{43} & A_{44} & 0 \\ A_{93} & A_{94} & A_{99} \end{pmatrix},$$

whose determinant is $\det(\mathcal{J}_{\text{red}} - \lambda I) = \det(\mathcal{A} - \lambda I) \cdot \det(\mathcal{C} - \lambda I)$. For the matrix $\mathcal{A}$, $Tr(A) = A_{11} + A_{22} = f_h'(N_h) - (\xi_h + 2\mu_h + \omega_p) < 0$ and $det(A) = A_{11}A_{22} - A_{12}A_{21} = -(\omega_p + \mu_h + \xi_h)(\mu_h - f_h'(N_h)) > 0$ since $\mu_h - f_h'(N_h) < 0$. Hence, the eigenvalues of $A$ are negative or have negative real parts.

The matrix $\mathcal{C}$ can be written in expanded form as

$$\mathcal{C} = \begin{pmatrix} A_{33} & 0 & A_{39} \\ A_{43} & A_{44} & 0 \\ A_{93} & A_{94} & A_{99} \end{pmatrix} = \begin{pmatrix} -(\gamma_h(M^*) + \delta_h(M^*) + \rho_h + \mu_h) & 0 & \frac{\beta(E^*,C^*)p_{vh}[S_h^* + (1-\varepsilon_p)V_h^*]}{N_h^*} \\ \gamma_h(M^*) & -(\omega_r + \mu_h) & 0 \\ \frac{\beta(E^*,C*)p_{hv}S_v^*}{N_h^*} & \frac{\beta(E^*,C^*)p_{hv}\theta S_v^*}{N_h^*} & -\mu_v \end{pmatrix}.$$

Observe that the block ($\mathcal{C}$) is associated with the infectious classes ($I_h, R_h$, and $I_v$) of the model (2.1), and that it can be written in the form $C = F - V$, where $F$ and $V$ are exactly the new infection and transition matrices used to determine the spectral radius $\rho(FV^{-1}) = \mathcal{R}_0^2$ through the next generation operator approach in Section S1.3. Hence, by the Next-Generation Matrix theorem [1], the eigenvalues of the block $\mathcal{C}$ satisfy $\text{Re}(\lambda) < 0 \iff \rho(FV^{-1}) < 1 \iff \mathcal{R}_0 < 1$. Therefore, the full disease-free equilibrium ($\mathcal{E}_{DFE}$) of the model (2.1) is LAS if $\mathcal{R}_0 < 1$ and unstable if $\mathcal{R}_0 > 1$.

*S1.5. Global stability of the disease-free equilibria*

*S1.5.1. Global stability of the trivial equilibrium ($\mathcal{E}_0$)*

We consider a linear Lyapunov function focused on the total populations $N_h$ and $N_v$: $L_1 = N_h + k_1 N_v$, where $k_1 > 0$ is a weighting constant. Taking the time derivative along the trajectories we obtain: $\dot{L}_1 = \dot{N}_h + k_1\dot{N}_v = [f_h(N_h) - \mu_h N_h - \delta_h(M)I_h] + k_1[f_v(N_v) - \mu_v N_v]$. Assuming the demographic functions satisfy $f_h(N_h) < \mu_h N_h$ and $f_v(N_v) < \mu_v N_v$ for all $N_h, N_v > 0$ (meaning the basic offspring numbers $\mathcal{T}_h$ and $\mathcal{T}_v$ are each less than 1): $\dot{L}_1 \leq (f_h(N_h) - \mu_h N_h) + k_1(f_v(N_v) - \mu_v N_v) \leq 0$. The derivative, $\dot{L}_1 = 0$ if and only if $N_h = 0$ and $N_v = 0$, which forces each of the variables $(S_h, V_h, I_h, R_h, S_v, I_v)$ to 0. On the set $\{N_h = 0,\ N_v = 0\}$, the remaining subsystem decouples: $\dot{M} = -\mu_h M$, $\dot{E} = \varepsilon_e E_0 - (\varepsilon_e + \sigma_e)E$, and $\dot{C} = -\sigma_c C$. Hence the only invariant subset is the trivial equilibrium $(0, 0, 0, 0, 0, E^*, 0, 0, 0)$. Substituting these into the remaining equations shows that the other variables also decay asymptotically to 0. By LaSalle's Invariance Principle, the trivial equilibrium is globally asymptotically stable (GAS) when $\mathcal{T}_h < 1$ and $\mathcal{T}_v < 1$.

*S1.5.2. Global stability of the mosquito-free equilibrium $\mathcal{E}_m$*

To prove the global stability of the mosquito-free equilibrium $\mathcal{E}_m = (S_h^*, V_h^*, 0, 0, M^*, E^*, 0, 0, 0)$, we construct a Lyapunov function focusing on the vector components: $L_2 = S_v + I_v$. Differentiating with respect to time gives: $\dot{L}_2 = \dot{S}_v + \dot{I}_v = f_v(N_v) - \mu_v(S_v + I_v) = f_v(N_v) - \mu_v N_v$. If the vector population cannot sustain itself ($\mathcal{T}_v < 1$), then $f_v(N_v) \leq \mu_v N_v$ for all $N_v$. Thus, $\dot{L}_2 \leq 0$.

The set where $\dot{L}_2 = 0$ is $\{N_v = 0\}$. On this invariant set, we have $S_v = I_v = 0$. Consequently, the ITN coverage equation becomes $\dot{C} = -\sigma_c C \Rightarrow C \to 0$, the ITN efficacy equation becomes $\dot{E} = \varepsilon_e(E_0 - E) - \sigma_e E \Rightarrow E \to E^*$, and the vector-to-human force of infection vanishes ($\lambda_{vh} = 0$). With $\lambda_{vh} = 0$, the infectious human and recovered compartments decay exponentially ($I_h \to 0,\ R_h \to 0$), and the human population reaches its demographic disease-free equilibrium ($S_h \to S_h^*$, $V_h \to V_h^*$). Finally, the immunity equation reduces to $\dot{M} = \tau_p \varepsilon_p V_h - \mu_h M$, implying $M \to M^*$.

Thus, the largest invariant subset of $\{\dot{L}_2 = 0\}$ is exactly the single equilibrium point $\mathcal{E}_m$. Therefore, by LaSalle's Invariance Principle, the system converges to $\mathcal{E}_m$ globally, i.e., $\mathcal{E}_m$ is globally asymptotically stable when $\mathcal{T}_v < 1$.

The largest invariant set where $\dot{L}_2 = 0$ is where $N_v = 0 \implies S_v = 0, I_v = 0$. When $I_v = 0$, the vector-to-human force of infection becomes $\lambda_{vh} = 0$. With $\lambda_{vh} = 0$, the human disease dynamics collapses, and $I_h \to 0, R_h \to 0$. Therefore, by LaSalle's Invariance Principle, the system converges to $\mathcal{E}_m$ globally, i.e., $\mathcal{E}_m$ is globally asymptotically stable when $\mathcal{T}_v < 1$.

*S1.5.3. Global stability of the full disease-free equilibrium ($\mathcal{E}_{DFE}$)*

To establish the global stability of the full disease-free equilibrium $\mathcal{E}_{DFE} = (S_h^*, V_h^*, 0, 0, M^*, E^*, C^*, S_v^*, 0)$, we use a linear combination of the human and vector infectious states: $L_3 = I_h + a_1 R_h + a_2 I_v$, where $a_1$ and $a_2$ are positive weighting constants to be determined. Differentiating $L_3$ along the system trajectories yields: $\dot{L}_3 = \dot{I}_h + a_1 \dot{R}_h + a_2 \dot{I}_v$. Letting $\tilde{A}_{33} = \gamma_h(M) + \delta_h(M) + \rho_h + \mu_h$, and substituting the expressions for $\dot{I}_h$, $\dot{R}_h$, and $\dot{I}_v$ leads to:

$$\dot{L}_3 = \lambda_{vh}[S_h + (1-\varepsilon_p)V_h] - \tilde{A}_{33} I_h + a_1\big[\gamma_h(M) I_h - (\omega_r + \mu_h) R_h\big] + a_2\big[\lambda_{hv} S_v - \mu_v I_v\big].$$

Using the definitions of $\lambda_{vh}$ and $\lambda_{hv}$, and noting that $S_h \le S_h^*$, $V_h \le V_h^*$, $S_h + (1-\varepsilon_p)V_h \le N_h$, $1/N_h \le 1/N_h^*$, and $S_v \le N_v \le N_v^*$:

$$\begin{aligned}\dot{L}_3 \quad &\le \quad \left(\frac{\beta(E^*, C^*) p_{vh} [S_h^* + (1-\varepsilon_p) V_h^*]}{N_h^*}\right) I_v - \tilde{A}_{33} I_h + a_1 \gamma_h I_h - a_1(\omega_r + \mu_h) R_h \\ &\quad + a_2 \left(\frac{\beta(E^*, C^*) p_{hv} N_v^*}{N_h^*}\right)(I_h + \theta R_h) - a_2 \mu_v I_v.\end{aligned}$$

To eliminate the explicit $R_h$ terms and group them with $I_h$, we match coefficients: $a_1(\omega_r + \mu_h) = a_2 \frac{\beta p_{hv} S_v^* \theta}{N_h^*} \implies a_1 = a_2 \frac{\beta(E^*, C^*) p_{hv} S_v^* \theta}{N_h^*(\omega_r + \mu_h)}$. Regrouping expression of $\dot{L}_3$ based on $I_h$ and $I_v$, we have :

$$\dot{L}_3 \le \left[a_2 \frac{\beta(E^*, C^*) p_{hv} S_v^*}{N_h^*}\left(1 + \frac{\theta \gamma_h}{\omega_r + \mu_h}\right) - \tilde{A}_{33}\right] I_h + \left[\frac{\beta_{max} p_{vh} [S_h^* + (1-\varepsilon_p) V_h^*]}{N_h^*} - a_2 \mu_v\right] I_v.$$

To clear the coefficient of $I_v$, we set: $a_2 = \frac{\beta(E^*, C^*) p_{vh} [S_h^* + (1-\varepsilon_p) V_h^*]}{\mu_v N_h^*}$. Substituting $a_2$ back into the expression and simplifying reveals the explicit expression for the threshold ($\mathcal{R}_0^2$):

$$\dot{L}_3 \le \tilde{A}_{33}\left[\frac{\beta(E^*, C^*)^2 p_{vh} p_{hv} S_v^* [S_h^* + (1-\varepsilon_p) V_h^*]}{\mu_v \tilde{A}_{33} (N_h^*)^2}\left(1 + \frac{\theta \gamma_h}{\omega_r + \mu_h}\right) - 1\right] I_h = \tilde{A}_{33}(\mathcal{R}_0^2 - 1) I_h.$$

If $\mathcal{R}_0^2 \le 1$, then $\dot{L}_3 \le 0$. The condition $\dot{L}_3 = 0$ holds strictly when $I_h = 0$. Direct substitution of $I_h = 0$ back into the model system forces $R_h \to 0$, which in turn interrupts vector transmission, forcing $I_v \to 0$. As a result, the remaining variables $(S_h, V_h, M, E, C, S_v)$ converge uniquely to their disease-free equilibrium values $(S_h^*, V_h^*, M^*, E^*, C^*, S_v^*)$. By LaSalle's Invariance Principle, the full disease-free equilibrium ($\mathcal{E}_{DFE}$) is globally asymptotically stable when $\mathcal{R}_0 \le 1$.

### S2. Examples of functional forms for the human and mosquito recruitment terms

Representative functional forms for human and mosquito recruitment terms $f_h$ and $f_v$ are presented in Table S1.

Table S1: Examples of human and mosquito recruitment terms.

| Pair | Human recruitment term | Mosquito recruitment term |
|---|---|---|
| 1 | Logistic recruitment: $f_h(N_h) = r_h N_h \left(1 - \frac{N_h}{K_h}\right)$ $r_h$ is the intrinsic human birth rate $K_h$ is the human carrying capacity | Logistic recruitment: $f_v(N_v) = r_v N_v \left(1 - \frac{N_v}{K_v}\right)$ $r_v$ is the intrinsic mosquito birth rate $K_v$ is the mosquito carrying capacity |
| 2 | Constant growth ($\delta_h = 0$): $f_h(N_h) = \mu_h N_h$ | Logistic recruitment: $f_v(N_v) = r_v N_v \left(1 - \frac{N_v}{K_v}\right)$ |
| 3 | Constant recruitment: $f_h(N_h) = \Lambda_h$ | Logistic recruitment: $f_v(N_v) = r_v N_v \left(1 - \frac{N_v}{K_v}\right)$ |
| 4 | Constant recruitment: $f_h(N_h) = \Lambda_h$ | Maynard-Smith growth: $f_v(N_v) = \frac{r_v N_v}{1+\left(\frac{N_v}{K_v}\right)^n}$, $n > 0$ |
| 5 | Constant recruitment: $f_h(N_h) = \Lambda_h$ | Constant recruitment: $f_v(N_v) = \Lambda_v$ |

We now demonstrate how to prove Theorem 3.1 using specific recruitment functions such as those presented in Table S1. Since the approach is similar, we prove only the first case with logistic human and logistic mosquito recruitment, i.e., the case for which $f_h = r_h N_h \left(1 - \frac{N_h}{K_h}\right)$ and $f_v = r_v N_v \left(1 - \frac{N_v}{K_v}\right)$, where $K_h(K_v)$ is the human (mosquito) carrying capacity and the fourth case with constant human recruitment (i.e., $f_h(N_h) = \Lambda_h$) and the Maynard-Smith-Slatkin mosquito recruitment term, $f_v(N_v) = (r_v N_v)/\left[1 + \left(\frac{N_v}{K_v}\right)^n\right]$, $n > 0$. A similar approach can be used for other recruitment function combinations.

*1. Logistic human and logistic mosquito recruitment.* Equations for the total human and mosquito populations are $\dot{N}_h = r_h N_h \left(1 - \frac{N_h}{K_h}\right) - \mu_h N_h - \delta_h(M) I_h$ and $\dot{N}_v = r_v N_v \left(1 - \frac{N_v}{K_v}\right) - \mu_v N_v$, which simplify to $\dot{N}_h \leq r_h N_h \left[\left(1 - \frac{N_h}{K_h}\right) - \mu_h\right] = (r_h - \mu_h) N_h \left(1 - \frac{N_h}{\tilde{K}_h}\right)$ and $\dot{N}_v = (r_v - \mu_v) N_v \left(1 - \frac{N_v}{\tilde{K}_v}\right)$, where $\tilde{K}_j = K_j(r_j - \mu_j)/r_j, j \in \{h, v\}$ are the new effective carrying capacities. Solving the differential inequality for $N_h$ with $N_h(0) = N_{h0}$, we obtain $N_h(t) \leq \frac{\tilde{K}_h N_{h0}}{N_{h0} + \left(\tilde{K}_h - N_{h0}\right) e^{-(r_h - \mu_h)t}} \to \tilde{K}_h$ as $t \to \infty$. Similarly, solving the differential equation for $N_v$ with $N_v(0) = N_{v0}$, we obtain $N_v(t) = \frac{\tilde{K}_v N_{v0}}{N_{v0} + \left(\tilde{K}_v - N_{v0}\right) e^{-(r_v - \mu_v)t}} \to \tilde{K}_v$ as $t \to \infty$. Combining this with the details and expressions for $E_{\max}, C_{\max}$ and $M_{\max}$ in Section S1.1, we conclude that the bounded region $\Omega_1 = \{(S_h, V_h, I_h, R_h, M, E, C, S_v, I_v) \in \mathbb{R}^9_+ : 0 \leq N_h \leq \tilde{K}_h, 0 \leq E \leq E_{\max}, 0 \leq C \leq C_{\max}, 0 \leq M \leq M_{\max},\ 0 \leq N_v \leq \tilde{K}_v\}$, is positively invariant and attracting.

*2. Constant human recruitment and Maynard-Smith-Slatkin mosquito recruitment.* The total human population is described by $\dot{N}_h = \Lambda_h - \mu_h N_h - \delta_h(M) I_h \leq \Lambda_h - \mu_h N_h$ since $\delta_h(M) \geq 0$ and $I_h(t) \geq 0$ for all $t \geq 0$. Solving this differential inequality with $N_h(0) = N_{h0}$ yields: $N_h(t) \leq \frac{\Lambda_h}{\mu_h} + \left(N_{h0} - \frac{\Lambda_h}{\mu_h}\right) e^{-\mu_h t} \to \frac{\Lambda_h}{\mu_h}$ as $t \to \infty$.

The total mosquito population is described by the equation $\dot{N}_v = \frac{r_v N_v}{1+\left(\frac{N_v}{K_v}\right)^n} - \mu_v N_v$. Assuming $r_v > \mu_v$ for population persistence, setting $\dot{N}_v = 0$ yields the unique positive steady-state carrying capacity $N_{v,\max} = K_v \left(\frac{r_v - \mu_v}{\mu_v}\right)^{1/n}$. If $N_v(t) > N_{v,\max}$, then $\dot{N}_v < 0$, and if $N_v(t) < N_{v,\max}$, then $\dot{N}_v > 0$. Thus, by standard comparison principles, $\limsup_{t \to \infty} N_v(t) = N_{v,\max}$.

Combining these with the details and expressions for $E_{\max}, C_{\max}$ and $M_{\max}$ in Section S1.1, while noting that $N_{h,max} = \Lambda_h/\mu_h$, we conclude that the bounded region $\Omega_4 = \{(S_h, V_h, I_h, R_h, M, E, C, S_v, I_v) \in \mathbb{R}^9_+ : 0 \leq N_h \leq \Lambda_h/\mu_h, 0 \leq E \leq E_{\max}, 0 \leq C \leq C_{\max}, 0 \leq M \leq M_{\max},\ 0 \leq N_v \leq N_{v,\max}\}$, is a globally attracting and positively invariant region for the system.

### S3. Existence and stability of disease-free equilibria for the model (2.1) with specific recruitment functions

We demonstrate the computation of the disease-free equilibrium of the model (2.1) for the specific example with constant human recruitment and Maynard-Smith-Slatkin mosquito recruitment. Computation of these equilibria for the other combinations of recruitment functions in Table S1 follows the same approach or simple substitutions of the functional forms for $f_h$ and $f_v$ in the general expressions derived in Section S1.2. It should be noted that there is no extinction equilibrium for the case of constant human recruitment and Maynard-Smith-Slatkin mosquito recruitment. Specifically, we have two equilibria, a mosquito-free equilibrium ($\mathcal{E}_m$) and a disease-free equilibrium ($\mathcal{E}_{DFE}$). From Section S1.2, the mosquito-free equilibrium is given by $\mathcal{E}_m = (S_h^*, V_h^*, 0, 0, M^*, E^*, 0, 0, 0)$, where $S_h^* = \frac{\omega_p+\mu_h}{\xi_h+\omega_p+\mu_h}N_h^*$, $V_h^* = \frac{\xi_h}{\xi_h+\omega_p+\mu_h}N_h^*$, $M^* = \frac{\tau_p\varepsilon_p\xi_h N_h^*}{\mu_h(\xi_h+\omega_p+\mu_h)+\omega_p\xi_h}$, $E^* = \frac{\varepsilon_e E_0}{\varepsilon_e+\sigma_e}$, and $N_h^*$ is the positive solution of the equation $f_h(N_h^*) = \mu_h N_h^*$. With $f_h(N_h^*) = \Lambda_h$, we have $\Lambda_h = \mu_h N_h^* \Rightarrow N_h^* = \Lambda_h/\mu_h$. Thus, $S_h^* = \frac{\Lambda_h(\omega_p+\mu_h)}{\mu_h(\xi_h+\omega_p+\mu_h)}$, $V_h^* = \frac{\Lambda_h\xi_h}{\mu_h(\xi_h+\omega_p+\mu_h)}$, $M^* = \frac{\Lambda_h\tau_p\varepsilon_p\xi_h N_h^*}{\mu_h[\mu_h(\xi_h+\omega_p+\mu_h)+\omega_p\xi_h]}$, and $E^* = \frac{\varepsilon_e E_0}{\varepsilon_e+\sigma_e}$.

From Section S1.2, the disease-free equilibrium is given by $\mathcal{E}_{DFE} = (S_h^*, V_h^*, 0, 0, M^*, E^*, C^*, S_v^*, 0)$ , where the non-zero state expressions are: $S_h^* = \frac{\omega_p+\mu_h}{\xi_h+\omega_p+\mu_h}N_h^*$, $V_h^* = \frac{\xi_h}{\xi_h+\omega_p+\mu_h}N_h^*$, $M^* = \frac{\tau_p\varepsilon_p\xi_h N_h^*}{\mu_h(\xi_h+\omega_p+\mu_h)+\omega_p\xi_h}$, $E^* = \frac{\varepsilon_e E_0}{\varepsilon_e+\sigma_e}$, $C^* = \frac{\varphi E^* N_v^* C_0}{\varphi E^* N_v^*+\sigma_c(1+\phi E^* N_v^*)}$, $S_v^* = N_v^*$, where $N_h^*$ is the positive solution to the host demographic equation: $f_h(N_h^*) = \mu_h N_h^*$ and $N_v^*$ is the positive solution of the equation: $f_v(N_v^*) = \mu_v N_v^*$. With $f_h(N_h^*) = \Lambda_h, N_h^* = \Lambda_h/\mu_h$. With $f_v(N_v) = \frac{r_v N_v}{1+\left(\frac{N_v}{K_v}\right)^n}$, we have $f_v(N_v) = \frac{r_v N_v}{1+\left(\frac{N_v}{K_v}\right)^n} = \mu_v N_v^*$. That is, $\frac{r_v N_v}{1+\left(\frac{N_v}{K_v}\right)^n} = [\mu_{v0} + \mu_{v1}E^*C^*]N_v^*$. This implies that $N_v^* = 0$ or $\frac{r_v}{1+\left(\frac{N_v}{K_v}\right)^n} = [\mu_{v0} + \mu_{v1}E^*C^*]$. Substituting the expressions of $E^*$ and $C^*$ and simplifying yields $\frac{r_v}{1+\left(\frac{N_v^*}{K_v}\right)^n} = \frac{\mu_{v0}\sigma_c+\left[\mu_{v0}(\varphi+\sigma_c\phi)E^*+\mu_{v1}\varphi(E^*)^2C_0\right]N_v^*}{\sigma_c+(\varphi+\sigma_c\phi)E^*N_v^*}$. With further simplification, this can be written as the following $(n+1)^{th}$ order polynomial equation in $N_v^*$: $p(N_v^*) = a_{n+1}(N_v^*)^{n+1} + a_n(N_v^*)^n - a_1 N_v^* - a_0 = 0$, where the explicit coefficients are: $a_{n+1} = \mu_{v0}(\varphi + \sigma_c\phi)E^* + \mu_{v1}\varphi(E^*)^2C_0 > 0$, $a_n = \mu_{v0}\sigma_c > 0$, $a_1 = K_v^n\left[(r_v - \mu_{v0})(\varphi + \sigma_c\phi)E^* - \mu_{v1}\varphi(E^*)^2C_0\right]$, $a_0 = K_v^n(r_v - \mu_{v0})\sigma_c > 0$ since $r_v > \mu_{v0}$. Observe that $a_{n+1}, a_n > 0$, and that $a_0 > 0$ if $r_v > \mu_{v0}$, which is required for mosquito persistence. Since $a_{n+1}, a_n, a_0 > 0$, there is exactly one sign change in the sequence of coefficients, irrespective of whether $a_1$ is positive or negative. Therefore, according to Descartes' Rule of Signs, the equation $p(N_v^*) = 0$ has at most one possible positive real solution ($N_v^* > 0$).

Substituting $\mathcal{E}_{DFE}$ in the expression for the reproduction number from Section S1.3, yields the specific reproduction number that corresponds to the constant human recruitment and Maynard-Smith-Slatkin mosquito recruitment functional forms. Also, substituting $\mathcal{E}_m$ and $\mathcal{E}_{DFE}$ in the Jacobian ($\mathcal{J}$) in Section S1.4 leads to the proof of the local stability of $\mathcal{E}_m$ and $\mathcal{E}_{DFE}$.

### S4. Data

Table S2: Annual malaria incidence data (2002–2022).

| Year | Incidence |
|---|---|
| 2002 | 20,049 |
| 2003 | 39,383 |
| 2004 | 28,328 |
| 2005 | 105,824 |
| 2006 | 149,584 |
| 2007 | 262,050 |
| 2008 | 839,903 |
| 2009 | 769,761 |
| 2010 | 898,531 |
| 2011 | 1,002,805 |
| 2012 | 1,453,471 |
| 2013 | 2,375,129 |
| 2014 | 2,851,555 |
| 2015 | 2,041,277 |
| 2016 | 3,064,796 |
| 2017 | 3,607,026 |
| 2018 | 2,318,090 |
| 2019 | 5,019,389 |
| 2020 | 4,069,277 |
| 2021 | 4,270,769 |
| 2022 | 4,890,691 |

### S5. Estimated parameters using the model with no vaccination

Initial conditions: The initial conditions were set as $S_h(0) = 32{,}609{,}759$, $V_h(0) = 0$, $I_h(0) = 55$, $R_h(0) = 100{,}000$, $M(0) = 150$, $E(0) = 0.65$, $C(0) = 0.1$, $S_v(0) = 163{,}139{,}375$, and $I_v(0) = 9{,}665$.

Table S3: Estimated parameter values with corresponding 95% bootstrap confidence intervals (CI) for the full malaria transmission model. Confidence intervals were obtained using 1000 bootstrap iterations. Model fitting yielded a sum of squared errors (SSE) of $7.29280 \times 10^{12}$, coefficient of determination $R^2 = 0.87530$, and optimizer exit flag $= 2$. Also shown are the estimated basic reproduction number ($\mathcal{R}_0$) and the disease-free equilibrium (DFE) state values: $S_h^* = 114348000$, $M^* = 0.00000$, $C^* = 0.84916$, $S_v^* = 292571559.51130$, $N_h^* = 114348000$, and $N_v^* = 292571559.51130$.

| Parameter | Estimate | 95% CI |
|---|---|---|
| $\eta$ | 0.36103 | [0.25512, 0.63323] |
| $\tau_d$ | 9.14608 | [0.69938, 9.79468] |
| $p_{vh}$ | 0.16850 | [0.15563, 0.19677] |
| $\delta_{h1}$ | 0.55421 | [0.04264, 0.95954] |
| $d$ | 352.14481 | [26.21695, 480.39060] |
| $\varphi$ | 1.49236 | [1.15552, 3.50562] |
| $\phi$ | 0.59568 | [0.38035, 1.11168] |
| $n$ | 2.14853 | [2.00111, 2.90247] |
| $\gamma_{h1}$ | 1.00679 | [0.14566, 8.66232] |
| $\alpha$ | 9.00609 | [7.64628, 11.03751] |
| $\beta_{\max}$ | 47.25630 | [44.83468, 51.54176] |
| $K_v$ | 524055280.57881 | [392327086.40695, 775383703.13143] |
| $r_v$ | 29.03946 | [23.91864, 31.43438] |
| $\mathcal{R}_0$ | 1.40307 | [1.27435, 1.49696] |

### S6. Supplementary figures

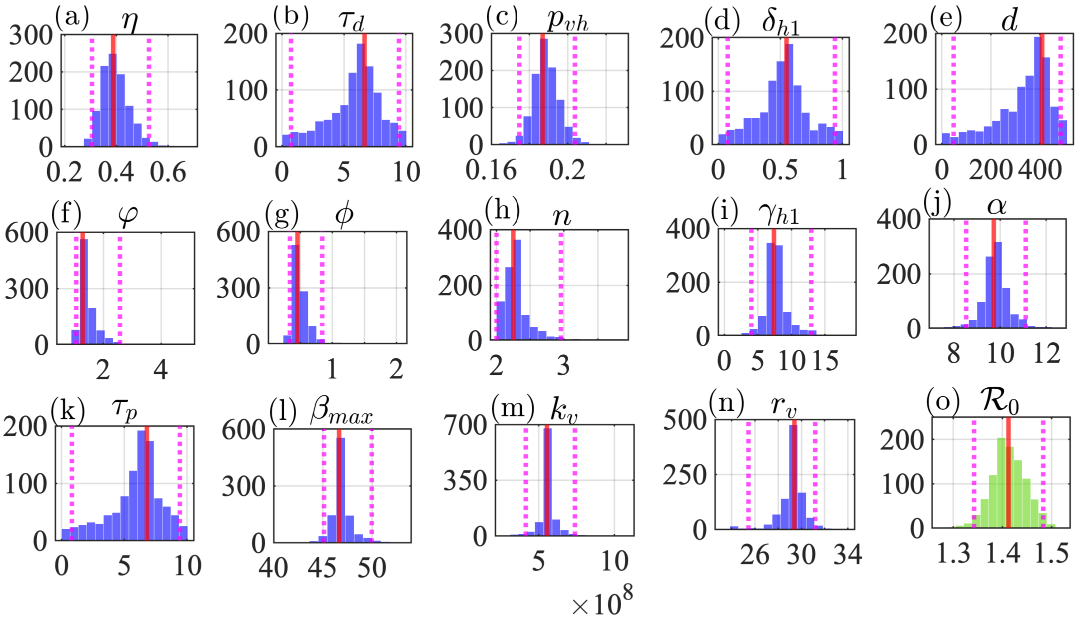


Figure S1: Parametric bootstrap distributions and 95% confidence intervals for estimated model parameter values and $\mathcal{R}_0$. Subplots 1–14 (blue histograms in (a)-(n)) display the empirical distributions of the model parameters, while subplot 15 (green histogram in (o)) displays the distribution for the Basic Reproduction Number ($\mathcal{R}_0$) derived via the Next-Generation Matrix method. For all 15-bin histograms, the horizontal axis denotes the estimated parameter range, and the vertical axis denotes the frequency count across 1000 bootstrap iterations. Solid red lines indicate the original point estimates obtained from the best fit to the empirical data, and dotted magenta vertical lines bound the 95% confidence intervals. The tight clustering of a distribution around its point estimate indicates high parameter certainty and strong model identifiability, whereas a widely spread or highly skewed distribution reveals increased parameter uncertainty; for subplot 15, a distribution situated entirely above 1.0 provides statistical verification of sustained transmission potential.

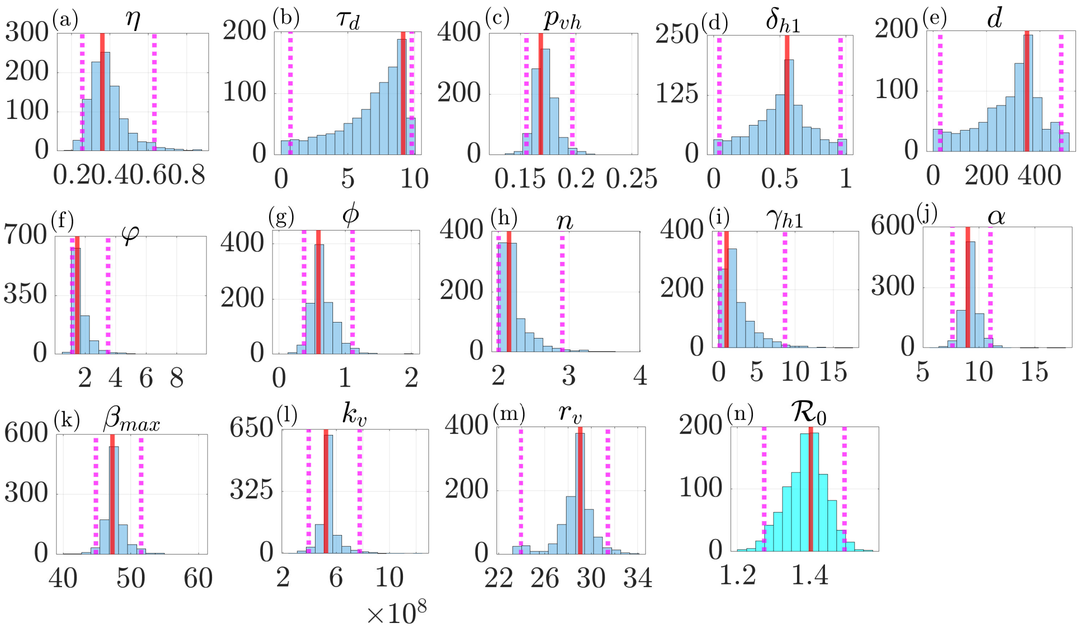


Figure S2: Parametric bootstrap distributions and 95% confidence intervals for estimated model parameter values and $\mathcal{R}_0$. Subplots 1–13 (histograms in (a)-(m)) display the empirical distributions of the model parameters, while subplot 14 (histogram in (n)) displays the distribution for the Basic Reproduction Number ($\mathcal{R}_0$) derived via the Next-Generation Matrix method. For all 15-bin histograms, the horizontal axis denotes the estimated parameter range, and the vertical axis denotes the frequency count across 1000 bootstrap iterations. Solid red lines indicate the original point estimates obtained from the best fit to the empirical data, and dotted magenta vertical lines bound the 95% confidence intervals. The tight clustering of a distribution around its point estimate indicates high parameter certainty and strong model identifiability, whereas a widely spread or highly skewed distribution reveals increased parameter uncertainty; for subplot 15, a distribution situated entirely above 1.0 provides statistical verification of sustained transmission potential.

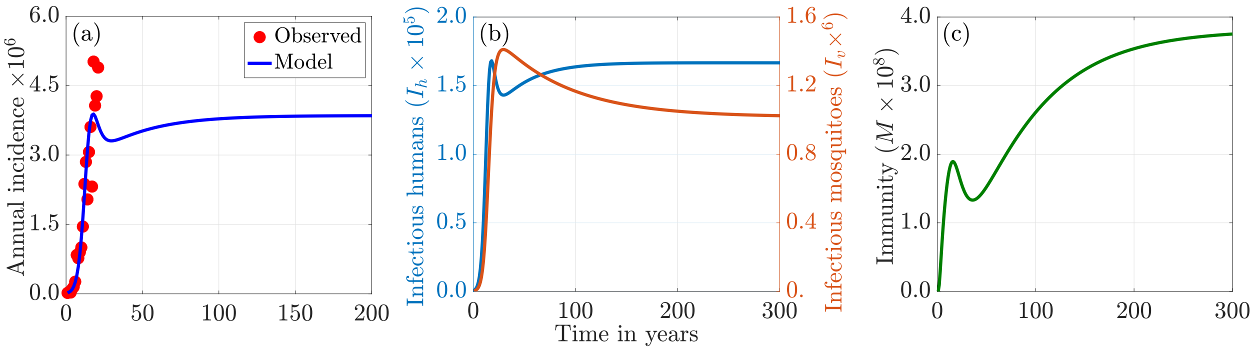


Figure S3: **Long-term dynamics of the model.** (a) Long-term incidence of newly reported cases over time; (b) infectious human and mosquito population dynamics, illustrating the simultaneous progression of active infections in both host ($I_h$) and vector ($I_v$) populations; and (c) population-level immunity dynamics, showing the evolution of the immune profile over an extended time horizon. The simulations were generated using the estimated and fixed parameter set in Table 2(b). The initial conditions used for this simulation are $S_{h0} = 32,609,759$, $V_{h0} = 0$, $I_{h0} = 55$, $R_{h0} = 100,000$, $M_0 = 150$, $E_0 = 0.65$, $C_0 = 0.1$, $S_{v0} = 163,139,375$, and $I_{v0} = 9,665$.

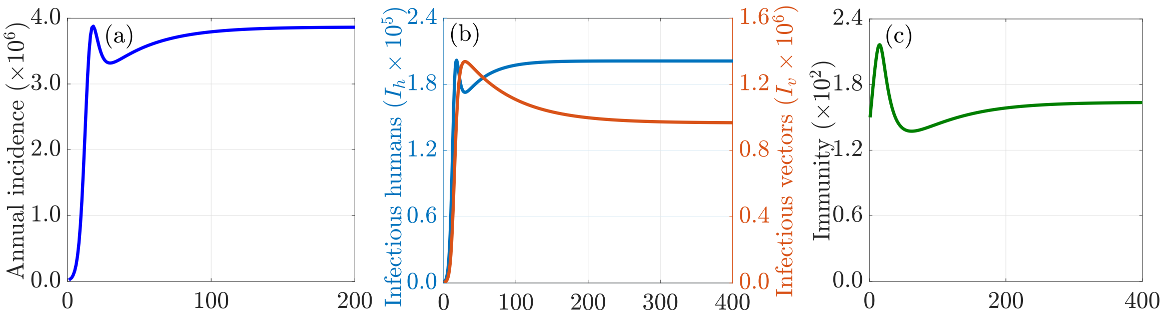


Figure S4: **Long-term dynamics of the model with no vaccination.** (a) Long-term incidence of newly reported cases over time; (b) infectious human and mosquito population dynamics, illustrating the simultaneous progression of active infections in both host ($I_h$) and vector ($I_v$) populations; and (c) population-level immunity dynamics, showing the evolution of the immune profile over an extended time horizon. The simulations were generated using the estimated and fixed parameter set in Table 2. The initial conditions used for this simulation are $S_{h0} = 32,609,759$, $I_{h0} = 55$, $R_{h0} = 100,000$, $M_0 = 150$, $E_0 = 0.65$, $C_0 = 0.1$, $S_{v0} = 163,139,375$, and $I_{v0} = 9,665$.

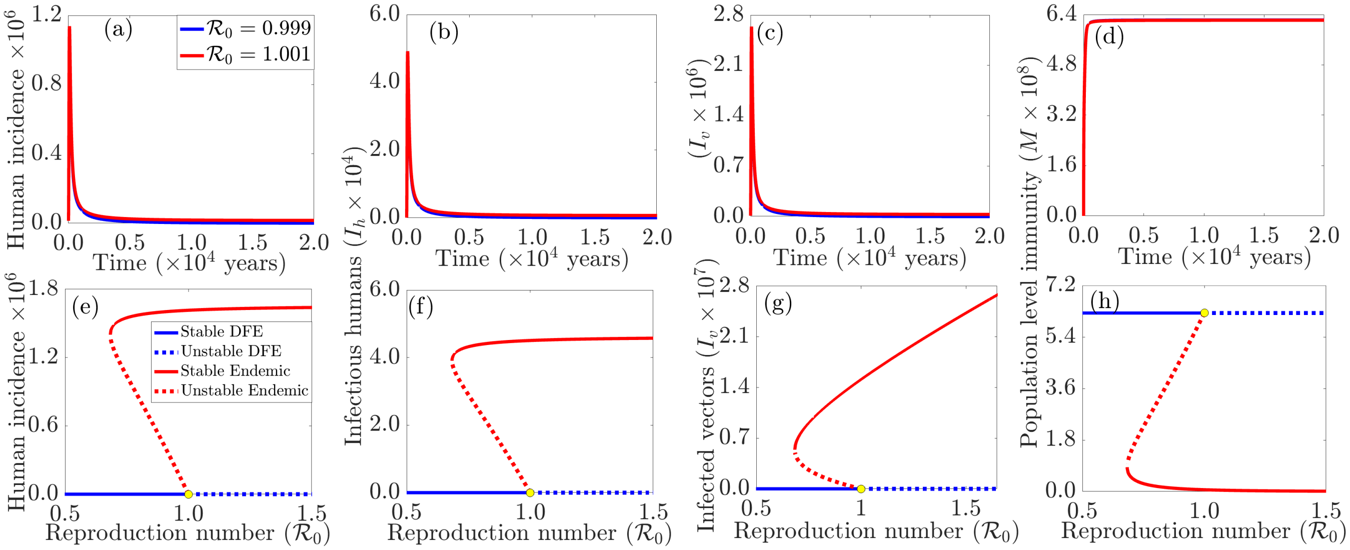


Figure S5: Long-term dynamics illustrating the threshold behaviour of $\mathcal{R}_0$ ((a)-(d)), and the emergence of backward bifurcation under a high malaria-induced mortality rate ((e)-(h)). Panels (a)-(d) show convergence to the disease-free equilibrium for $\mathcal{R}_0 = 0.999$ and to an endemic equilibrium for $\mathcal{R}_0 = 1.001$, confirming the classical threshold property of $\mathcal{R}_0$. Panels (e)-(h) show the coexistence of stable disease-free and endemic equilibria when the malaria-induced mortality rate is increased to 1000 times its baseline value, demonstrating the occurrence of backward bifurcation. The simulations were initiated in 2002 using the initial conditions $S_h(0) = 32{,}609{,}759$, $V_h(0) = 0$, $I_h(0) = 55$, $R_h(0) = 100{,}000$, $M(0) = 150$, $E(0) = 0.65$, $C(0) = 0.1$, $S_v(0) = 163{,}139{,}375$, and $I_v(0) = 9{,}665$, together with the parameter values listed in Tables 2 of the main text.

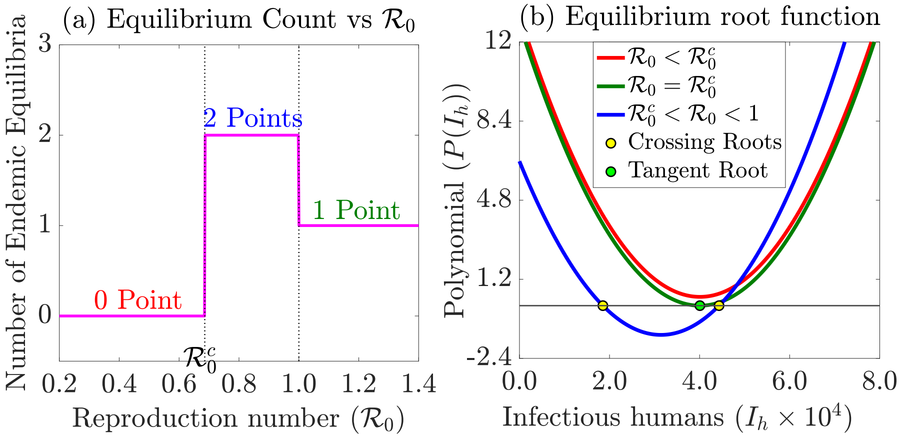


Figure S6: Bifurcation analysis and number of endemic equilibrium points as a function of the basic reproduction number $\mathcal{R}_0$. (a) The exact count of endemic equilibrium points plotted against $\mathcal{R}_0$. This profile highlights the regions of parameter space where the system has no endemic equilibria when $\mathcal{R}_0$ is below a critical threshold ($\mathcal{R}_0^c$), a unique endemic equilibrium, or two endemic equilibria (indicative of backward bifurcation or bistability). (b) Algebraic determination of the endemic states via the equilibrium polynomial $P(I_h^*)$ plotted against the infectious human population at equilibrium ($I_h^*$). The real, positive roots of this polynomial determine the existence and exact number of endemic equilibria. Specifically, the number of endemic equilibria corresponds to the number of times the polynomial curve cuts the horizontal ($I_h^*$) axis (where $P(I_h^*) = 0$) within the biologically feasible domain $I_h^* \in (0, N_h]$.
